# The Sex-Specific Role of Adrenal Androgens in Youth Psychopathology

**DOI:** 10.1101/2025.09.30.25336972

**Authors:** Franka Edith Weisner, Bianca Serio, Sofie Valk, Luise Bläschke, Franziska Degenhardt, Anke Hinney, Raphael Hirtz, Lars Dinkelbach

## Abstract

Adolescence is a vulnerable period for the emergence of mental health problems. Adrenarche, an early stage of pubertal development marked by rising adrenal androgens, particularly dehydroepiandrosterone (DHEA), may influence emotional and behavioral development. However, longitudinal evidence linking preadolescent endocrine influences to adolescent psychopathology remains limited. Using data from the Adolescent Brain Cognitive Development (ABCD) Study (N_max_=10,562), we analyzed whether salivary DHEA during preadolescence predicted later externalizing and internalizing symptoms during adolescence. To create a more robust estimate for hormonal levels in preadolescence, hormone concentrations were averaged across baseline and 1-year follow-up (age range=8.9-12.4 years). Outcomes were measured via the Child Behavior Checklist (CBCL) at the 2-, 3-, and 4-year follow-ups (age range=10.6-15.8 years). Sex-stratified linear mixed models were employed, adjusting for age, race/ethnicity, BMI and physical activity. In males, higher DHEA levels were linked to fewer externalizing symptoms across all follow-ups (e.g., β=–0.07 SD change of CBCL per SD-change of log-transformed DHEA levels (95% CI [–0.10, –0.04] at 3-year) and to fewer internalizing symptoms at 3-year and 4-year follow-ups. The effect of preadolescent DHEA in males translated into a reduced probability of externalizing symptoms crossing borderline or clinical thresholds at each follow-up (e.g., adjusted Risk Ratio=0.76 to reach clinical threshold for CBCL externalizing per SD increase in log-transformed DHEA; 95% CI [0.62, 0.98] at 3-year). In females, no hormone–symptom associations emerged. Interestingly, sex-by-DHEA interaction effects increased with age for both symptom domains. These findings suggest that preadolescent adrenal endocrine influences may play a role in thedevelopment of sex-specific vulnerability during adolescence. Future studies should consider adrenarche as a sensitive period for hormonal effects on mental health.

## Background

Adolescence marks a critical developmental window during which many mental health problems begin to emerge^1,2^. This period is also characterized by a notable shift in mental health vulnerability between the sexes^3–6^. During childhood, males typically exhibit higher levels of psychiatric symptoms, especially in the form of externalizing behaviors such as aggression and hyperactivity^3^. As adolescence begins, females show a pronounced rise of internalizing symptoms, most notably depression and anxiety^3,4^. These internalizing problems become the most prevalent form of psychopathology in adolescence overall^5^, while externalizing problems in boys tend to persist but represent a smaller part of the overall burden^5,6^.

The transitional period from childhood to adolescence, defined here as preadolescence spanning ages 9 to 12, is increasingly recognized as a critical period for brain maturation and emotional development^7–10^. From an endocrine perspective, preadolescence involves the onset of two distinct but overlapping maturational processes: gonadarche and adrenarche. Although closely interrelated, gonadarche and adrenarche are governed by distinct control mechanisms and can occur independently of one another^11–13^. Gonadarche involves activation of the hypothalamic-pituitary-gonadal (HPG) axis, leading to the production of gonadal testosterone and estradiol and the visible physical changes of puberty, including breast development in females, testicular growth in males, pubic and axillary hair growth and accelerated height gain in both sexes^14^. This process is widely considered the hormonal hallmark of puberty and both the timing and progression of these physical changes have been repeatedly linked to mental health outcomes^11,12,15–17^. In contrast, adrenarche precedes puberty and marks the endocrine transition from childhood to adolescence^18,19^. It is an often underrecognized phase of pubertal development, characterized by increased secretion of androgens, particularly dehydroepiandrosterone (DHEA) and its sulfated form (DHEA-S), from the adrenal glands^13,14,18^. Within in the central nervous system, DHEA and DHEA-S have been shown to be negative noncompetitive modulators of GABA_A_ receptors, to potentiate glutaminergic N-methyl-D-aspartate (NMDA) receptors, and to increase neurite differentiation and neurogenesis, suggesting a potential role in neurodevelopment^20–22^. Fluctuations in adrenal androgens during preadolescence may influence neural circuits involved in emotion regulation, stress reactivity, and social behavior – processes that undergo substantial refinement during that period of development and are critical for mental health vulnerability^8,23–25^.

Building on this neurobiological rationale for a potential link between endocrine transitions and psychopathological symptoms, several studies have investigated associations between DHEA and youth psychopathology. These studies have been dominated by cross-sectional designs^26–32^, with only limited longitudinal investigations^25,33–35^. Some studies report positive associations between DHEA levels and aggression or conduct problems^26,28,29^, while DHEA-S levels have also been linked to fewer externalizing symptoms in adolescent males^36^. For internalizing problems, both positive and negative associations between DHEA levels and symptom severity have been reported^25,26,29,30,32^. Other studies did not detect consistent associations between DHEA or DHEA-S levels and psychopathologies^27,33,37^.

Overall, the literature on DHEA is conflicting and points to heterogeneous and often sex-specific associations, with effect directions varying by developmental stage, outcome domain, and study design^16,27,37–39^. A recent systematic review found that current evidence does not permit firm conclusions regarding associations between androgens and adolescent mental health and identified several methodological weaknesses likely contributing to the mixed findings^37^. First, many studies linking higher androgen levels to mental health risks rely on high-risk or clinical samples, creating potential selection and/or collider bias. Second, longitudinal designs remain scarce, limiting the ability to draw inference on the direction and causality of reported associations. Third, inappropriate adjustment for confounders was common. Additionally, few studies have explicitly targeted preadolescence^25–28^, leaving the role of preadolescent hormone exposure (particularly adrenarche) underexplored, despite proposals that it represents a sensitive period for neurodevelopment, mental health, and the emergence of sex differences^7–9^.

Together, these inconsistencies underscore the need for large-scale, longitudinal, and sex-stratified investigations of how hormones in preadolescence may be associated with psychopathology at later stages of puberty. To address these gaps, we analyzed data from the longitudinal, population-based Adolescent Brain Cognitive Development (ABCD) Study^40^ to examine whether preadolescent hormone profiles predict externalizing and internalizing psychopathology in later adolescence. Our analyses focus on levels and changes of DHEA and are complemented by analyses of testosterone and estradiol. By analyzing sex-specific patterns in endocrine markers, we sought to clarify the role of hormones in shaping sex-specific mental health trajectories across adolescence.

## Methods

### Sample Description and Exclusion Criteria

This study analyzed data from the Adolescent Brain Cognitive Development (ABCD) Study, a large longitudinal, population-based cohort comprising 11,868 children and adolescents recruited across 21 sites in the United States^40^. The ABCD Study conducts annual assessments, including the collection of hormonal saliva samples. For this analysis, we used release 5.1 of the dataset (DOI: 10.15154/z563-zd24), accessed via the National Institute of Mental Health Data Archive. The release includes data from baseline (N=11,868, mean age=9.91±0.62 years, 47.8% female), 1-year (N=11,220, mean age=10.92±0.64 years, 47.7% female), 2-year (N=10,973, mean age=12.03±0.67 years, 47.5% female), 3-year (N=10,336, mean age=12.91±0.65 years, 47.5% female) and 4-year follow-ups (N=4,754, mean age=14.08±0.68 years, 47.6% female).

Data quality was assessed at two levels (Supplementary Figure S1). At the subject level, entire participants were excluded if they had missing or implausible key demographic data (missing age N=1, missing race/ethnicity N=1, missing or implausible BMI values N=60), reported congenital differences in sex development (N=3), or completely lacked hormonal measurements at both baseline or 1-year follow-up (N=1,232), rendering them unable to contribute to the current study. At the observation level, specific time points were excluded as part of hormone-specific quality control procedures (e.g., refusal of saliva sample, sex mismatch, or technical assay concerns). Additionally, estradiol was assessed only in females per ABCD protocol^41^.

These criteria resulted in 49,151 included observations from up to N=10,562 (89.0%) participants (Table 1), where each observation corresponds to a participant’s measurement at a single time point. Across all follow-up years, excluded participants did not differ in age compared to included participants but were more likely to be of black or hispanic backgrounds and more frequently came from lower-income families (Supplementary Tables S1–S3).

**Table 1.**
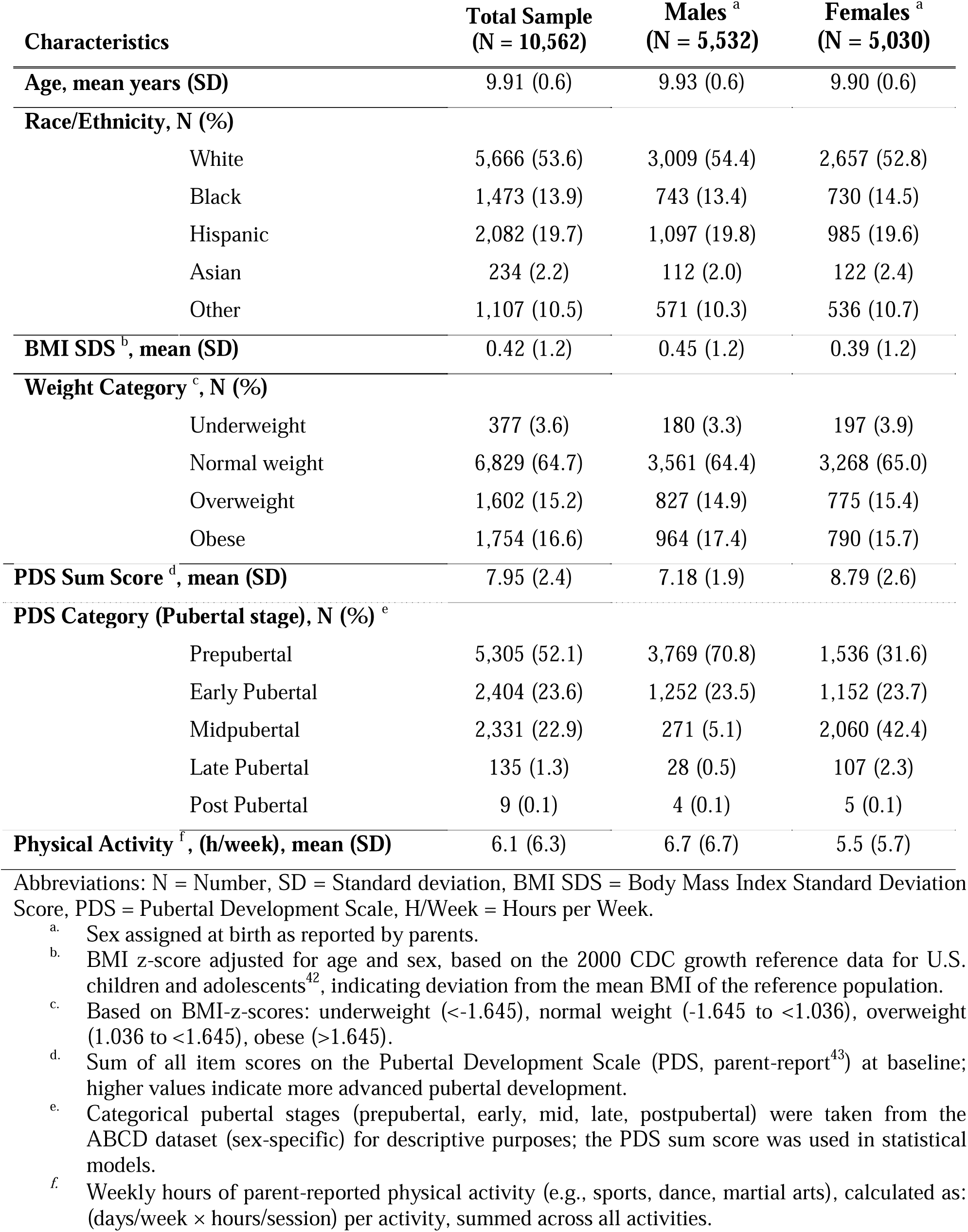
Baseline Characteristics of the Total and Sex-Stratified Sample.

| Characteristics | Total Sample<br>(N = 10,562) | Males<sup>a</sup><br>(N = 5,532) | Females<sup>a</sup><br>(N = 5,030) |
| --- | --- | --- | --- |
| Age, mean years (SD) | 9.91 (0.6) | 9.93 (0.6) | 9.90 (0.6) |
| Race/Ethnicity, N (%) |  |  |  |
| White | 5,666 (53.6) | 3,009 (54.4) | 2,657 (52.8) |
| Black | 1,473 (13.9) | 743 (13.4) | 730 (14.5) |
| Hispanic | 2,082 (19.7) | 1,097 (19.8) | 985 (19.6) |
| Asian | 234 (2.2) | 112 (2.0) | 122 (2.4) |
| Other | 1,107 (10.5) | 571 (10.3) | 536 (10.7) |
| BMI SDS<sup>b</sup>, mean (SD) | 0.42 (1.2) | 0.45 (1.2) | 0.39 (1.2) |
| Weight Category<sup>c</sup>, N (%) |  |  |  |
| Underweight | 377 (3.6) | 180 (3.3) | 197 (3.9) |
| Normal weight | 6,829 (64.7) | 3,561 (64.4) | 3,268 (65.0) |
| Overweight | 1,602 (15.2) | 827 (14.9) | 775 (15.4) |
| Obese | 1,754 (16.6) | 964 (17.4) | 790 (15.7) |
| PDS Sum Score<sup>d</sup>, mean (SD) | 7.95 (2.4) | 7.18 (1.9) | 8.79 (2.6) |
| PDS Category (Pubertal stage), N (%)<sup>e</sup> |  |  |  |
| Prepubertal | 5,305 (52.1) | 3,769 (70.8) | 1,536 (31.6) |
| Early Pubertal | 2,404 (23.6) | 1,252 (23.5) | 1,152 (23.7) |
| Midpubertal | 2,331 (22.9) | 271 (5.1) | 2,060 (42.4) |
| Late Pubertal | 135 (1.3) | 28 (0.5) | 107 (2.3) |
| Post Pubertal | 9 (0.1) | 4 (0.1) | 5 (0.1) |
| Physical Activity<sup>f</sup>, (h/week), mean (SD) | 6.1 (6.3) | 6.7 (6.7) | 5.5 (5.7) |

Abbreviations: N = Number, SD = Standard deviation, BMI SDS = Body Mass Index Standard Deviation Score, PDS = Pubertal Development Scale, H/Week = Hours per Week.
<sup>a</sup>. Sex assigned at birth as reported by parents.
<sup>b</sup>. BMI z-score adjusted for age and sex, based on the 2000 CDC growth reference data for U.S. children and adolescents<sup>42</sup>, indicating deviation from the mean BMI of the reference population.
<sup>c</sup>. Based on BMI-z-scores: underweight (<-1.645), normal weight (-1.645 to <1.036), overweight (1.036 to <1.645), obese (>1.645).
<sup>d</sup>. Sum of all item scores on the Pubertal Development Scale (PDS, parent-report<sup>43</sup>) at baseline; higher values indicate more advanced pubertal development.
<sup>e</sup>. Categorical pubertal stages (prepubertal, early, mid, late, postpubertal) were taken from the ABCD dataset (sex-specific) for descriptive purposes; the PDS sum score was used in statistical models.
<sup>f</sup>. Weekly hours of parent-reported physical activity (e.g., sports, dance, martial arts), calculated as: (days/week × hours/session) per activity, summed across all activities.

### Ethics

Written informed consent was obtained from legal guardians and assent from all participating children. The ABCD protocol was approved by the central institutional review board.at UC San Diego and, at some sites, additionally by local institutional review boards. Data use for this study was approved by the Ethics Committee of the Medical Faculty, University of Duisburg-Essen (24-12040-BO).

### Exposure and Outcome

To examine whether early hormonal profiles predict later mental health outcomes, we used hormone levels (averaged across baseline and 1-year follow-up) as well as hormone changes (ratio of baseline to 2-year follow-up) to predict CBCL scores assessed at the 2-, 3-, and 4-year follow-ups.

### Exposure: Hormone levels and changes

As part of the ABCD study protocol, salivary levels of DHEA, testosterone, and estradiol (females only) were assayed using the passive drool method and analyzed via Salimetrics enzyme immunoassay kits^41^. The dataset, including hormone concentrations, was obtained from the National Institute of Mental Health Data Archive. At each time point, each hormone sample was assayed in two technical replicates. The final hormone value per time point was calculated as the arithmetic mean of both replicates; if only one replicate was available, that single value was used. As raw hormone concentrations were heavily right skewed, log-transformation of the mean hormone concentrations was performed to approximate a normal distribution (see Supplementary Figures S2-S3).

Values falling below the lower limit of detection (LLD; DHEA: <5pg/ml, N=629; testosterone: <1pg/ml, N=9; estradiol: <0.1pg/ml, N=225) were imputed on the log-transformed scale using truncated normal regression models. Imputation, rather than exclusion of values below the LLD, was applied to retain information on low hormone concentrations. Imputations were conducted separately for each hormone and time point. Sex and age were included as predictors for imputation to retain biological plausibility. Imputed values remained below the respective LLD and approximated a truncated normal distribution consistent with the expected distribution of log-transformed hormone levels (see Supplementary Figures S6-S11).

The final hormone values were then used to derive preadolescent hormone levels and changes: To estimate hormonal levels throughout preadolescence and to create a more robust estimate, we computed the hormone level as the arithmetic mean of the log-transformed baseline (age 9.91±0.62 years) and 1-year follow-up (age 10.92±0.64 years) values for each hormone and participant. If only baseline (N=1,052) or 1-year follow-up (N=1,465) was available, that single time point was used. To capture expected changes in hormone concentrations associated with age and pubertal progression, a hormone change variable was calculated. This variable is defined as the natural logarithm of the ratio of the 2-year follow-up (age 12.03±0.67 years) value to the baseline hormone value. Our analyses focus on DHEA, which serves as a biomarker of adrenarche and reflects the predominance of adrenal androgens during this developmental phase^8,13^. In addition, we conducted exploratory analyses including estradiol and testosterone as further pubertal hormones and related sex steroids.

### Outcome: Child Behavior Checklist (CBCL)

Psychopathology was assessed annually using the parent-report Child Behavior Checklist (CBCL), a standardized questionnaire for evaluating emotional and behavioral problems in children and adolescents over the past six months. The 113 items are rated on a three-point Likert scale and are used to compute summary scores for Total, Internalizing, and Externalizing Problems, with higher scores reflecting greater difficulties^44^.

To assess whether early hormonal profiles predicted later mental health outcomes, CBCL data from the 2-year, 3-year, and 4-year follow-up assessments served as outcome variables in longitudinal analyses. For primary analyses, we used the raw scores of the syndrome-oriented CBCL scales, namely “CBCL Externalizing” (comprising Rule-Breaking and Aggressive Behavior) and “CBCL Internalizing” (comprising Anxious/Depressed, Withdrawn/Depressed, and Somatic Complaints). CBCL data availability decreased across follow-ups, with nearly complete coverage up to 3 years (DHEA data: 3-year N=8,914). For the 4-year follow-up, only about half of the sample was included in release 5.1, resulting in CBCL availability of N=4,085 (Supplementary Figure S1).

### Covariates

To identify relevant confounders, we used the framework of Directed Acyclic Graphs (DAG, constructed in DAGitty.net^45^, Supplementary Figure S5). This approach allowed us to systematically consider relationships not only between exposure, covariates, and outcome, but also among potential covariates. Based on this framework, the following minimally sufficient adjustment set to adjust for potential confounding was identified: age at baseline (continuous), parent-reported race/ethnicity (five-category factor), BMI standard deviation score at baseline and physical activity at baseline (for rationale of covariate selection, see Supplemental Material and S5).

### Statistical Analysis

After evaluating the assumptions of linear modeling (see Supplementary Table S5-S7, Supplementary Figure S12–S14), we estimated separate linear mixed-effects models (LMMs) for each hormone and outcome, including the above-mentioned set of covariates and a random intercept for study site and a random intercept family nested within study site to account for clustering effects. All hormone and outcome variables were standardized, so that the resulting beta estimates represent changes in the outcome in standard deviations per one standard deviation change in the exposure. All models were stratified by sex, given possible differences in hormonal systems between males and females and the study’s focus on sex-specific effects. In addition, we tested for sex-by-DHEA interaction effects to assess whether associations between DHEA levels and mental health outcomes differed by sex across follow-up years. To explore the potential clinical relevance of the results, two further analyses were conducted: First, we estimated adjusted risk ratios (RRs) for the association between preadolescent hormone levels (averaged across baseline and 1-year follow up) or changes (the ratio of 2-year follow-up to baseline values) with CBCL defined thresholds of internalizing and externalizing problems at 2-, 3-, and 4-year follow-ups. Risk ratios were obtained from binomial regression models with a log link, adjusted for the same covariates as the main analyses and stratified by sex. CBCL outcomes were categorized into borderline (T-score≥60) and clinical (T-score≥64) ranges. A detailed description of this analysis is provided in the Supplementary Results (Risk Ratio Analysis). Second, we explored associations between DHEA levels and changes within the six DSM-5-oriented CBCL subscales (Depression, Anxiety, Attention-Deficit/Hyperactivity Disorder (ADHD), Oppositional Defiant Disorder (ODD), Conduct Disorder (CD), and Somatic Problems)^46^, which more closely align with clinical diagnostic categories.

### Sensitivity and Secondary Analyses

To assess the robustness of our main findings, we conducted several sensitivity analyses. Given that the CBCL outcome scores represent count data with a right-skewed distribution (Supplementary Figure S4) and that QQ-plots indicated a violation of the assumption of the normal distribution of residuals (Supplementary Figure S12 and S13), we re-estimated all models using negative binomial mixed models. To account for potential confounding by psychotropic or hormone-related medication use, we repeated analyses after excluding participants who reported taking such medications at baseline. Additional sensitivity analyses were performed adjusting for pubertal status at baseline and at the 3-year follow-up to account for changes in pubertal development over time. To address potential bias from imputation of hormone values below the lower limit of detection, we conducted analyses excluding participants with hormone values below the lower limit of detection. Lastly, accounting for prior symptomatology (i.e., higher CBCL scores at baseline predicting higher scores at later time points), an expanded model included baseline CBCL scores as an additional covariate. As secondary analyses, we repeated all models using preadolescent testosterone and estradiol levels (females only) as exposures.

## Results

We tested whether preadolescent DHEA levels levels (averaged across baseline and 1-year follow up) or changes (the ratio of 2-year follow-up to baseline values) predicted later psychopathology by modeling associations with CBCL outcomes across follow-ups. All associations were further examined by testing sex-by-DHEA interaction effects, translating results into risk ratios for developing CBCL scores in the borderline or clinical range for each respective outcome, and examining DSM-5–oriented CBCL subscales. Primary analyses focused on the relationship between DHEA and CBCL Externalizing and Internalizing scores at the 3-year follow-up. Further, the investigation was expanded to include testosterone and estradiol (females only) and the 2-year and 4-year follow-up, assessing consistency across hormones and time points.

### The Effect of DHEA on Psychopathology

Preadolescent DHEA levels (mean age males 10.41±0.64 years, mean age females 10.38±0.64 years) differed between sexes, with females exhibiting higher salivary DHEA levels than males (males=57.67±40.44 pg/ml; females=77.94±54.91 pg/ml; t-test: <0.001).

### DHEA and CBCL Outcomes at 3-Year Follow-Up

At 3-year follow up higher DHEA levels in males were associated with lower externalizing symptoms scores (Figure 1). This association remained after adjusting for covariates, showing an estimated effect size of β=–0.07 SD change of CBCL per SD-change of log-transformed DHEA levels (95% CI [–0.10, –0.04]). The association with internalizing symptoms in males was weaker, with an estimated effect size of ß=-0.03, 95% CI [-0.06, 0.00]. In females, DHEA levels were neither associated with externalizing (β=0.00, 95% CI [–0.03, 0.03]) nor with internalizing symptoms (β=0.03, 95% CI [0.00, 0.06]). Changes in DHEA throughout preadolescence, calculated as the 2-year follow-up to baseline ratio, did not predict CBCL outcomes in either sex. Effect estimates for the association between DHEA change and CBCL symptom scales were small, and all confidence intervals included zero (adjusted models: males – externalizing: β=0.01, 95% CI [–0.02, 0.05]; internalizing: β=0.00, 95% CI [–0.04, 0.04]; females – externalizing: β=0.00, 95% CI [–0.04, 0.04]; internalizing: β=–0.02, 95% CI [–0.06, 0.02]).

**Figure 1.**
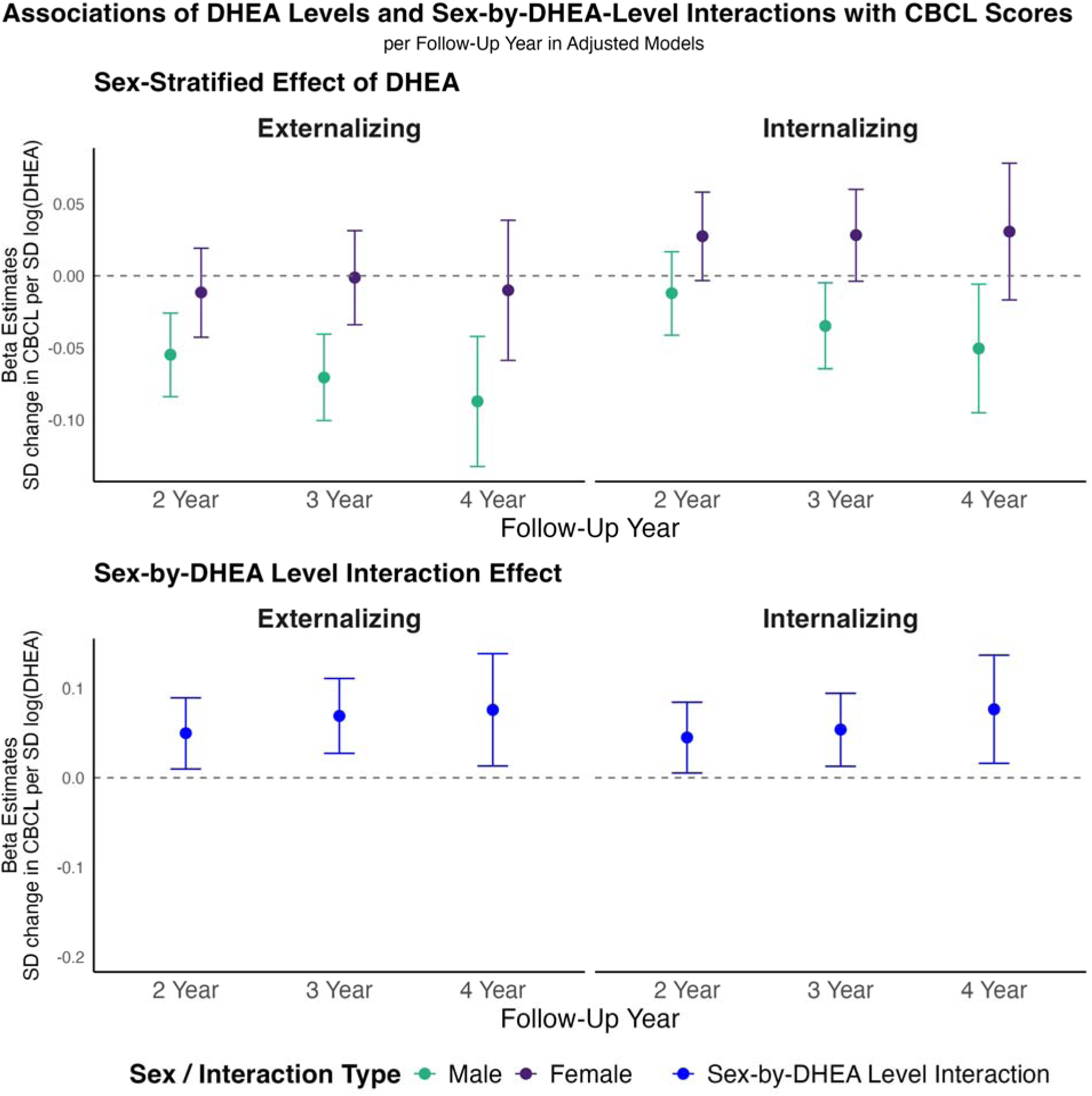
Sex-stratified associations and sex-by-DHEA Level interaction effects of DHEA on Child Behavior Checklist (CBCL) scores across follow-up years, derived from linear mixed models. DHEA level is defined as the arithmetic mean of the log-transformed baseline and 1-year follow-up DHEA values per participant. Panels display standardized beta estimates for the association between DHEA levels and CBCL outcomes (Externalizing and Internalizing scores) at 2-, 3-, and 4-year follow-up. Top row: Sex-stratified effects of DHEA for males (teal) and females (purple). Bottom row: Sex-by-DHEA interaction effects (blue) indicate the differential association according to sex. The y-axis represents standardized beta estimates (standard deviation change in CBCL score per standard deviation change in log-transformed DHEA level); error bars denote 95% CIs. Models are adjusted for age, race/ethnicity, BMI SDS, and physical activity, all assessed at baseline. Sample sizes for the 2-year, 3-year, and 4-year follow-ups for both Externalizing and Internalizing scores are N=5,029 male and N=4,578 female participants; N=4,676 male and N=4,238 female participants; and N=2,147 male and N=1,938 female participants, respectively. Abbreviations: 95%-CI = 95% Confidence Interval; BMI SDS=Body Mass Index Standard Deviation Score; CBCL=Child Behavior Checklist; DHEA=Dehydroepiandrosterone; N=Number; SD=Standard Deviation.

### Longitudinal Stability of DHEA Effects

To test the longitudinal stability of our findings and investigate potential developmental shifts, we extended our analyses to 2- and 4-year follow-ups (Figure 1). The negative association between DHEA levels and later externalizing symptoms in males increased from β=–0.05 (95% CI [–0.08, –0.03]) at 2-year follow-up to β=–0.09 (95% CI [–0.13, –0.04]) at 4-year follow-up, although the effect sizes remains small. Internalizing symptoms in males also showed negative associations at later follow-ups (2-year follow-up: β=–0.01, 95% CI [–0.04, 0.02]; 4-year follow-up: β=–0.05, 95% CI [–0.09, 0.00]). In females, associations for both externalizing and internalizing symptoms were minimal and fluctuated around zero across all follow-ups.

### Sex-by-DHEA Interaction Effects

For DHEA levels, interaction estimates for both externalizing and internalizing symptoms showed an increasing trend over time, with increasing sex × DHEA level interaction effects in later adolescence (Figure 1). For externalizing symptoms, interaction estimates increased from β=0.05 (95% CI [0.00, 0.09]) at 2-year follow-up to β=0.08 (95% CI [0.01, 0.14]) at 4-year follow-up. A similar pattern was observed for internalizing symptoms (2-year follow-up: β=0.04, 95% CI [0.00, 0.08]; 4-year follow-up: β=0.08, 95% CI [0.02, 0.14]). All sex × DHEA change interaction terms remained non-significant (see Supplementary Table S19).

### Risk for Symptoms Reaching Clinical Thresholds

The effects of DHEA levels on CBCL scores translated into differences in the risk of exceeding established thresholds for borderline (T-score≥60) and clinical (T-score≥64) levels of psychopathological symptoms (Figure 2). In males, higher DHEA levels were linked to a reduced risk to reach borderline or clinical-levels of externalizing symptom severity (T-score≥60) across all follow-up years: The adjusted risk ratio per SD-change of log-transformed DHEA levels was 0.84 (95% CI [0.74, 0.95]) at 2-year follow-up and further declined to 0.73 (95% CI [0.58, 0.92]) at 4-year follow-up. The strongest effect emerged at the clinical cutoff (T≥64) at 4-year follow-up: RR=0.55 (95% CI [0.40, 0.77]). There was no clear evidence for a relationship between DHEA levels and internalizing problems in males, nor for either symptom domain in females.

**Figure 2.**
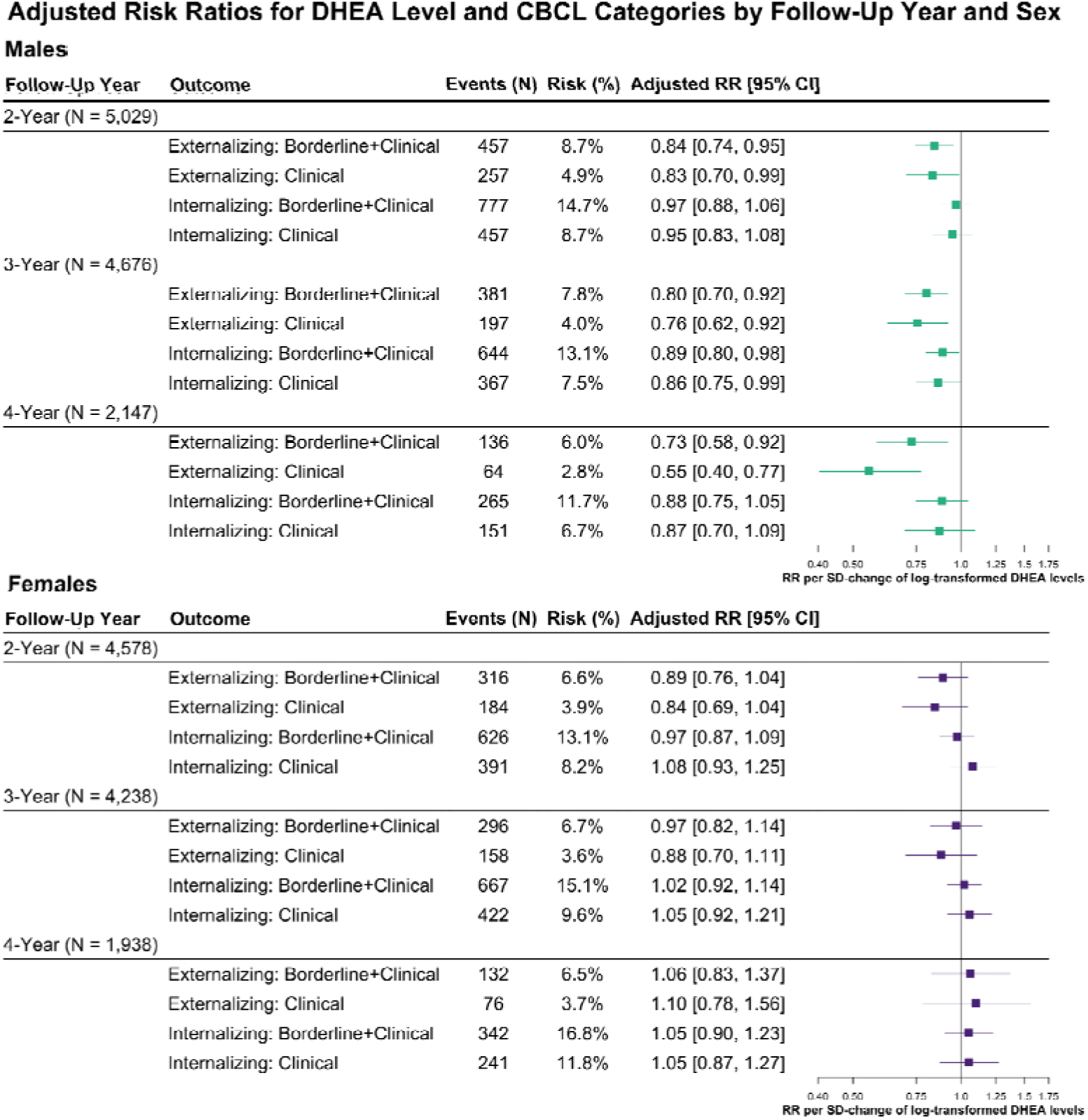
Sex-stratified risk ratios for the associations between DHEA levels and Child Behavior Checklist (CBCL) diagnostic categories (Externalizing and Internalizing, each divided into Borderline+Clinical and Clinical) across 2-, 3-, and 4-year follow-ups. Adjusted risk ratios (RRs) were estimated using linear mixed models. DHEA level is defined as the arithmetic mean of the log-transformed baseline and 1-year follow-up DHEA values per participant. Panels present adjusted RRs and 95% CIs for the associations between DHEA levels and CBCL outcomes separately for males (top panel, teal) and females (bottom panel, purple). The y-axis lists CBCL outcome categories; the x-axis shows adjusted RRs per SD-change of log-transformed DHEA levels. Squares represent point estimates; horizontal bars indicate 95% CIs. CBCL Score Interpretation: Normal Range: T-score≤59; Borderline+Clinical Range: T-score≥60; Clinical Range: T-score≥64. Events (N) indicates the number of participants meeting the CBCL threshold at the respective follow-up. Risk (%) represents the proportion of participants in that category relative to the total number with available data. Models are adjusted for age, race/ethnicity, BMI SDS, and physical activity, all assessed at baseline. Sample sizes for the 2-year, 3-year, and 4-year follow-ups for both Externalizing and Internalizing scores are N=5,029 male and N=4,578 female participants; N=4,676 male and N=4,238 female participants; and N=2,147 male and N=1,938 female participants, respectively. Abbreviations: 95% CI=95% Confidence Interval; BMI SDS=Body Mass Index Standard Deviation Score; CBCL=Child Behavior Checklist; DHEA=Dehydroepiandrosterone; N=Number; RR=Risk Ratio; SD=Standard Deviation.

### DHEA and CBCL Subdomains

Associations between DHEA levels and DSM-5-oriented subscales were primarily evident in males (Supplementary Figures S16-18). Higher DHEA levels were linked to lower symptoms of Conduct Problems, Oppositional Defiant Problems, and ADHD across all follow-up years. In females, there was no consistent pattern across subscales, though a transient association with lower ADHD symptoms was observed at the 2-year follow-up. For internalizing symptom subscales, negative associations between DHEA levels and Anxiety and Depression Problems were observed in males at the 3- and 4-year follow-ups, although effect sizes were smaller than those for externalizing symptoms.

### Effects of Testosterone and Estradiol on Psychopathology

Analyses revealed strong positive correlations between DHEA levels and both testosterone (males: mean r=0.75, females: mean r=0.80) and estradiol levels (females only, mean r=0.55, see Figure 3 and Supplementary Table S18) across all timepoints and remained high after controlling for pubertal status. Strong correlations indicate that multicollinearity prevents statistically disentangling the independent effects of DHEA, testosterone, and estradiol.

**Figure 3.**
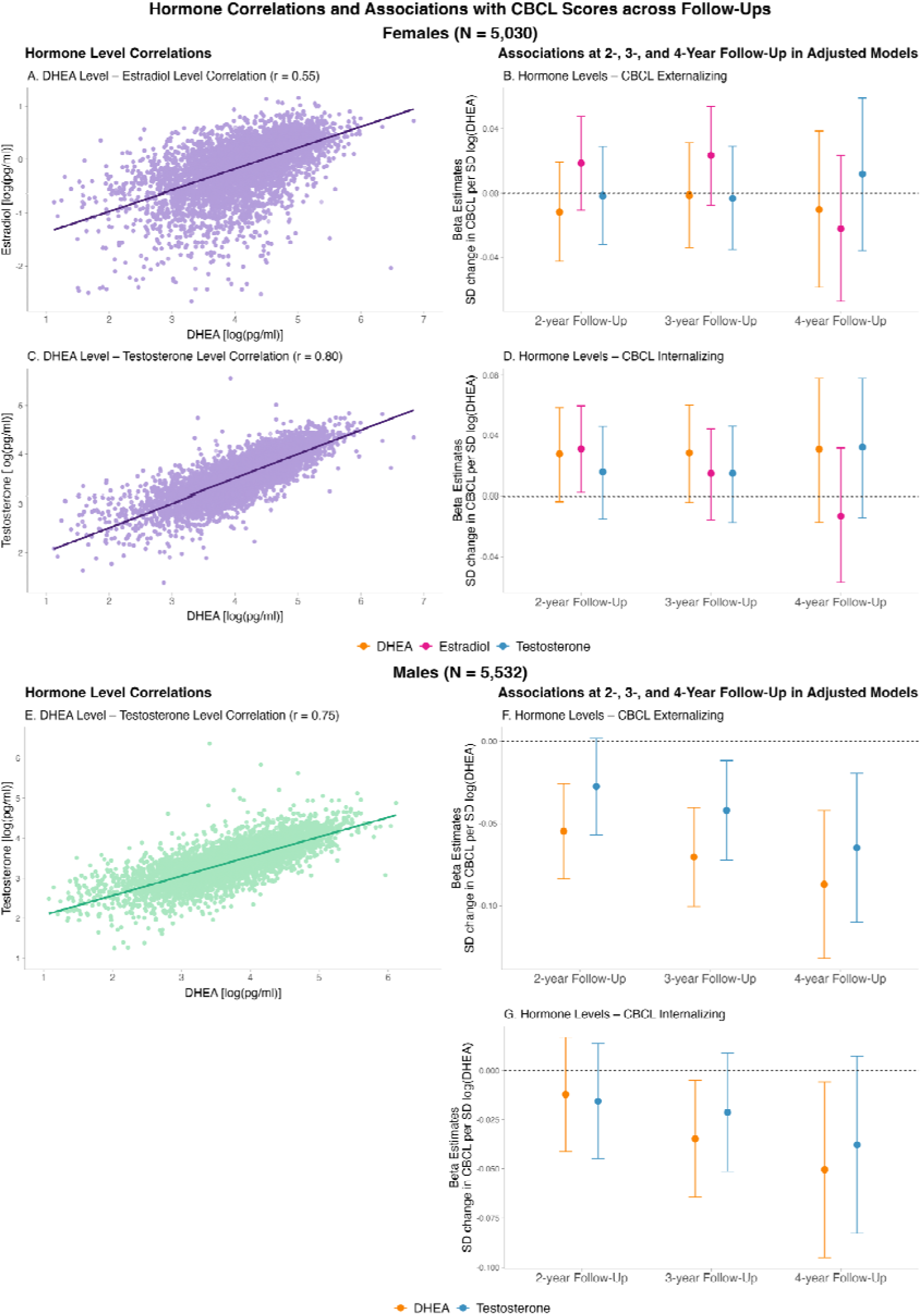
Sex-stratified hormone level correlations and associations between hormone levels and CBCL scores across 2-, 3-, and 4-year follow-ups. Panels A–D show results for females (N=5,030): A, C: Pairwise correlations between DHEA and Estradiol (A) and DHEA and Testosterone (C) with linear regression lines and Pearson’s r. B, D: Beta estimates (points) and 95% CIs (error bars) for the associations between hormone levels (DHEA, Estradiol, Testosterone) and CBCL Externalizing (B) and Internalizing (D) scores at 2-, 3-, and 4-year follow-up. Beta estimates reflect SD change of CBCL per SD-change of log-transformed hormone level. Panels E–G show results for males (N=5,532): E: Pairwise correlation between DHEA and Testosterone with a linear regression line and Pearson’s r. F, G: Beta estimates (points) and 95% Cis (error bars) for the associations between hormone levels (DHEA, Testosterone) and CBCL Externalizing (F) and Internalizing (G) scores at 2-, 3-, and 4-year follow-up. Colors: orange: DHEA; pink: Estradiol; blue: Testosterone. DHEA is re-shown from Figure 1 for comparison to the effects of testosterone and estradiol. Hormone levels were defined as the arithmetic mean of the log-transformed baseline and 1-year follow-up hormone values per participant [log(pg/ml)]. All models are adjusted for age, race/ethnicity, BMI SDS and physical activity, all assessed at baseline. Abbreviations: 95%-CI = 95% Confidence Interval; BMI SDS=Body Mass Index Standard Deviation Score; CBCL=Child Behavior Checklist; DHEA=dehydroepiandrosterone; r=Pearson’s R Correlation Coefficient; SD=Standard Deviation.

In males, effects of preadolescent testosterone levels (averaged across baseline and 1-year follow-up) on CBCL domains showed a similar pattern to the effects of DHEA levels: Testosterone levels showed consistent negative associations with externalizing symptoms and smaller effect estimates with internalizing symptoms across all timepoints (Figure 3). Confidence intervals were generally wider for testosterone than for DHEA. In females, none of the three hormones showed consistent associations with CBCL outcomes. Effect estimates varied across time points, and all confidence intervals included zero (Figure 3).

To test whether testosterone provided additional explanatory power beyond adrenal DHEA, we included testosterone as an additional exposure in the adjusted 3-year mixed-effects models. Adding testosterone did not meaningfully increase the proportion of variance explained in CBCL externalizing or internalizing scores in either males or females (see Supplementary Figure S15 for model statistics).

### Sensitivity Analyses

To test the robustness of our findings, we conducted several sensitivity analyses addressing methodological and biological concerns. Re-estimating models using negative binomial regression to account for the skewed distribution of CBCL scores (Supplementary Tables S8–S9), excluding participants taking psychotropic or hormone-related medication at baseline (Supplementary Tables S10-S11), adjusting for pubertal status at baseline (Supplementary Tables S12-S13) and at 3-year follow-up (Supplementary Tables S14-S15), and excluding rather than imputing hormone values below the detection threshold (Supplementary Tables S16-S17) all produced results similar to the primary findings. Controlling for baseline CBCL scores slightly attenuated the effect estimates of DHEA levels on externalizing CBCL scores in males, although the associations remained significant. The sex-by-DHEA interaction estimates remained largely unchanged when including baseline CBCL scores as a covariate in the model (see Supplementary Figure S15 and Supplementary Table S19).

## Discussion

This longitudinal study investigated whether DEHA levels in preadolescence predicted later externalizing and internalizing symptoms in a large population-based cohort of youth (N_max_=10,562). To capture hormone levels throughout preadolescence (age 9-12 years) and create a more robust estimate, hormone concentrations were averaged between log-transformed baseline and 1-year values. Outcomes were assessed up to four years later (mean age males 14.09±0.69 years, mean age females 14.07±0.67).

### Preadolescent DHEA-levels are linked to lower Externalizing and Internalizing Symptoms in Males

Our findings indicate that higher preadolescent DHEA levels are associated with lower externalizing symptom severity in males. This association was consistent across all externalizing symptom subscales (Conduct, Oppositional Defiant, and ADHD Problems) and across all follow-up assessments. Although effect sizes were small, they extended to a reduced likelihood to develop borderline or clinical-level symptoms. Similar, but less consistent effects were observed for internalizing problems.

Our findings contrast with studies using clinical cohorts that have linked higher adrenal androgen levels to clinically diagnosed externalizing problems in the context of conduct disorder^28,47,48^. Such associations have also been discussed in a systematic review concluding that adrenal androgens may serve as risk markers for psychopathology^8^. However, clinical samples are prone to selection bias, and their findings may not generalize to population-based cohorts^37^. Currently existing population-based studies have yielded mixed results^25–27,29–31,37^. One cross-sectional study found a negative association between DHEA-S and self-reported externalizing symptoms in 226 boys aged 11–12 years^36^, whereas a longitudinal study of 56 boys aged 10–14 years reported positive associations between DHEA/DHEA-S and symptoms of depression and anxiety^30^. Others found no such associations^27,37^. The heterogeneity in existing findings likely reflects methodological variation, as outlined by a recent systematic review^37^, particularly in sample composition, hormonal assessment, and covariate selection. In the current study, we leveraged population-based data, averaged hormone concentrations over time to create more robust hormone values, and examined the longitudinal influence of DHEA on mental health outcomes while adjusting for age, race/ethnicity, BMI, and physical activity. These design choices not only address methodological limitations of prior research^37^, such as reliance on high-risk or clinical samples, scarcity of longitudinal designs, and insufficient control of confounding factors, but also extend the field conceptually by focusing on preadolescent hormone levels as predictors of later psychopathology.

Our findings support models proposing that the transition from childhood to adolescence, endocrinologically marked by adrenarche, represents a sensitive window for neurodevelopment^7–9^. Although gonadarche has often been emphasized as a key pubertal milestone^14^, our results suggest that adrenarche, as an earlier endocrine transition, may also play a role in shaping mental health trajectories. DHEA, a hormonal surrogate marker of adrenarche^13,18^, may influence behavior via central neurophysiological pathways. As a neurosteroid, DHEA modulates GABAergic and glutamatergic signaling, alters cortical excitability, and shapes neurodevelopment by promoting neurite outgrowth and neurogenesis^18,20,22^. Adrenarche-related hormonal shifts are thought to influence frontolimbic brain systems involved in impulse control and executive function^49,50^, which are closely related to externalizing psychopathology^51,52^, including regions such as the prefrontal cortex and amygdala^25,53,54^. Future studies are needed to clarify the neuropathophysiological mechanisms underlying the current findings and specifically should address the sex-specific role of adrenal androgens for frontolimbic neurodevelopment in preadolescent boys.

### Preadolescent DHEA-levels have Sex-Specific Effects on Adolescent Mental Health

Adolescence marks a notable shift in the prevalence of mental health problems between sexes, with boys showing more externalizing symptoms during childhood and internalizing symptoms increasing sharply during adolescence in females^3,4^, becoming the most prevalent form of psychopathology in adolescence among sexes^5^. Our findings revealed DHEA × sex interaction effects across adolescence, indicating that the strength and direction of the association between DHEA and psychopathology differ by sex. Specifically, higher DHEA levels were associated with psychopathologic symptom burden only in boys. The divergence between the DHEA effects on mental health in boys and girls were robust to several sensitivity analyses and tended to increase over time, suggesting that sex differences in DHEA effects become more pronounced as adolescence progresses.

Our findings contribute to a body of research suggesting that associations between adrenal androgens and mental health outcomes may be moderated by sex. Consistent with our findings, several studies reported null associations in girls^27,33,55,56^ and some reported substantially stronger associations in boys compared to girls^29–31^. It has been proposed that sex differences may not emerge until later in adolescence^27^, a hypothesis consistent with our observation that interaction effects between sex and hormone levels became more pronounced over time. The current study emphasizes the idea of sex differences in hormone–psychopathology associations and further supports the idea that boys and girls differ in their neuroendocrine sensitivity and developmental trajectories.

Del Giudice et al (2009) proposed that preadolescence (or juvenile transition) is a neurodevelopmental and psychosocial sensitive period for the development of sex differences in neuroendocrine regulation and behavior. During this period, sex differences in aggression^57,58^ and attachment patterns^59^ become more apparent. Furthermore, genetic and environmental contributions to behavior vary by both age and sex, becoming pronounced in middle childhood, with stronger genetic effects in boys^60,61^. Those neurodevelopmental changes are thought to be, at least in part, mediated by adrenarche^7^, which initiates hormonal cascades promoting sexual differentiation in the brain prior to gonadal puberty^62,63^. More recent evidence links adrenal androgens such as DHEA to cortical maturation and spontaneous neural activity in preadolescents, indicating that adrenarche actively shapes neurodevelopmental pathways relevant to emotion and cognition^53,64^. Additionally, interactive effects among adrenal androgens (DHEA, testosterone) and further with stress hormones (e.g., cortisol) have been reported to change during adrenarche and adolescence, impacting brain and behavioral development and contributing to sex-divergent trajectories^53,65,66^.

Taken together, these findings strengthen the view that preadolescence constitutes a sensitive window with high adaptive plasticity^7,8^, during which adrenal androgens may act as mediators of neurodevelopmental divergence between boys and girls. Extended follow-up studies are needed to assess whether the here found sex-specific and progressively increasing effects of DHEA on youth psychopathology persist and how they contribute to sex differences in adult mental health.

### Effects of Testosterone and Estradiol May Reflect Shared Adrenal Origin

In our study, testosterone mirrored DHEA effects and showed comparable associations with externalizing and internalizing symptoms in males (Figure 3), that remained consistent when adjusting for pubertal status in a sensitivity analysis. DHEA and its sulfate ester (DHEA-S) are produced by the adrenal glands and are considered hallmark biomarkers of adrenal androgen production and adrenarche^18^. In contrast, estradiol and testosterone are typically viewed as gonadal pubertal hormones, whose production increases during gonadarche via activation of the hypothalamic–pituitary–gonadal axis^14^. However, DHEA and DHEA-S also serve as precursors for more potent sex steroids, including testosterone and estradiol, through peripheral (including gonadal) enzymatic conversion, making them part of a complex and interconnected steroid hormone network^18,23,67,68^. Because adrenarche precedes gonadarche, adrenal-derived pathways are supposed to provide the main source of circulating testosterone and estradiol in late childhood and the early phase of adolescence^23,67,68^. gonadal contributions to circulating testosterone and estradiol are expected to be minimal during preadolescence^18^, consistent with evidence that gonadal steroids increase later than adrenal ones^69^. Consistent with a presumably shared adrenal origin, we observed high correlations of DHEA levels with testosterone and estradiol, even after adjustment for the current pubertal stage (Supplementary Table S18). The effects of preadolescent testosterone and estradiol on later CBCL externalizing and internalizing scores largely mirrored those of preadolescent CBCL scores, and adding testosterone alongside DHEA did not meaningfully increase the variance explained in either outcome (Supplementary Results). Although we cannot definitively determine the source of circulating testosterone and estradiol in our sample, the findings suggest that their associations with psychopathology in preadolescence likely reflect shared adrenal activity rather than independent gonadal effects.

Recent studies on the contribution of steroids to adolescent mental health emphasize the importance of viewing steroid hormones as interdependent components within a shared metabolic architecture^70,71^. Examining hormonal constellations have been proposed as more robust predictors of mental health outcomes than single hormone values and allows to describe specific hormonal profiles for psychopathologies^70,71^. However, the description of these hormonal profiles relies on cost-intensive techniques such as gas or liquid chromatography–mass spectrometry (GC-/LC-MS)^72^. Although GC/LC-MS has been used to link adrenal dysregulation with depression in a clinical sample of adolescents^70,73^, population-based studies employing these methods are still needed. Future studies should employ steroid profiling with GC/LC-MS to simultaneously capture related steroid levels and thereby disentangle adrenal and gonadal contributions to the mental health effects of steroid hormones in population-based samples.

### Limitations and Future Directions

Our study has several limitations. First, our findings relied on parent-reported symptom scores from the CBCL. Although widely used to assess psychopathology symptom burden in children and adolescents, the CBCL has recently been subject to critical debate^74^. Originally designed as a screening tool to detect clinical levels of psychopathology in individuals, it was not intended to measure latent psychopathology traits in population-based samples, and its proposed dimensional structure failed to replicate in the ABCD cohort^74^. Recent attempts to develop an improved factor structure have also been unsuccessful^74^, and no similarly comprehensive alternative measures are available in the ABCD dataset. Given that the limitations of the CBCL are primarily expected to attenuate associations between biological variables and psychopathology^74^ (i.e., increase the likelihood of false negatives), we do not expect these limitations to compromise the validity of our current findings.

Second, we were not able to account for all contextual factors that may influence the relationship between adrenal androgens and mental health. Several studies have reported such interactions, for example, modulation of the association between DHEA and internalizing symptoms by salivary alpha-amylase concentrations^36^, effects dependent on the DHEA response to stressful situations^75^, or alterations in diurnal rhythmicity patterns^23,35^.

Third, hormone levels were measured via salivary samples using immunoassay (ELISA) methods, which, although widely applied in developmental research, have lower specificity than mass spectrometry–based approaches as GC/LC-MS. Future studies may employ these techniques which also allow to disentangle adrenal and gonadal hormone sources, identify pathway-specific markers, and better understand the role of single hormones on brain development^70,71^.

### Conclusions

Our findings add to a growing, yet heterogeneous, body of research on hormonal influences during the transition from late childhood to early adolescence. Negative associations between preadolescent DHEA levels and externalizing symptoms in males suggest a potential protective role of adrenal androgens during preadolescence and were translated to a reduced risk to reach borderline or clinical-levels of symptom severity. The longitudinal increase in hormone– psychopathology associations, together with emerging sex-by-DHEA interactions, points to a potential role of adrenal androgens in the sex-specific developmental shift of mental health problems across puberty and highlight adrenarche as a neurodevelopmental window influencing the emergence of psychiatric symptoms in adolescence.

## Data Availability

This study used data from the Adolescent Brain Cognitive Development (ABCD) Study, a project of the U.S. National Institutes of Health. ABCD data are available under controlled access via the National Institute of Mental Health Data Archive (NDA). Researchers can apply for access through the NDA data access process at https://nda.nih.gov/abcd, subject to approval to ensure compliance with ethical guidelines and participant confidentiality.

https://abcdstudy.org/

https://nda.nih.gov/abcd

## Abbreviations

95%-CI: 95% Confidence Interval
ABCD Study: Adolescent Brain Cognitive Development Study
ADHD: Attention Deficit Hyperactivity Disorder
BMI: Body Mass Index
BMI SDS: Body Mass Index Standard Deviation Score
CBCL: Child Behavior Checklist
DAG: Directed Acyclic Graph
DHEA: Dehydroepiandrosterone
DHEA-S: Dehydroepiandrosterone-sulfate
DSM-5: Diagnostic and Statistical Manual of Mental Disorders, Fifth Edition
ELISA: Enzyme-Linked Immunosorbent Assay
GC-/LC-MS: Gas Chromatography or Liquid Chromatography–Mass Spectrometry
HPG axis: Hypothalamic-Pituitary-Gonadal Axis
LLD: Lower Limit of Detection
LMM: Linear Mixed-Effects Models
N: Number
NIH: National Institutes of Health
PDS: Pubertal Development Scale
RR: Risk Ratio
SD: Standard Deviation.

## Author contributions

LD, RH, BS and AH conceptualized the study. Statistical analysis and data curation were performed by FEW. The original draft was written by FEW. LD supervised the statistical analyses, and reviewed and edited the manuscript. All authors (BS, SV, LB, AH, RH, FD) critically revised and approved the final manuscript.

## Funding

This work was supported by a fellowship from the University Medicine Essen Clinician Scientist Academy (UMEA) of the Medical Faculty of the University Duisburg-Essen (supported by the German Research Foundation (DFG)) to LD (FU 356/12-2).

## Data sharing

This study used data from the Adolescent Brain Cognitive Development (ABCD) Study, a National Institutes of Health (NIH) research project^76^. ABCD data are available via the National Institute of Mental Health Data Archive (NDA) under controlled access. Researchers can request data access via the NDA data application process (https://nda.nih.gov/abcd). Data access is granted after approval to ensure compliance with ethical guidelines and participant confidentiality. For further information, please visit the official ABCD Study^77^ or NDA websites^78^.

## Competing interests

FEW, BS, SV, LB, AH, RH, and LD have nothing to disclose. FD is supported by the BMFTR in the framework of the BMFTR Advanced Clinician Scientist Programme UMEA; 01EO2104.

## Acknowledgments

Data used in the preparation of this article were obtained from the Adolescent Brain Cognitive DevelopmentSM (ABCD) Study (Protected link to abcdstudy.org), held in the NIMH Data Archive (NDA). This is a multisite, longitudinal study designed to recruit more than 10,000 children age 9-10 and follow them over 10 years into early adulthood. The ABCD Study® is supported by the National Institutes of Health and additional federal partners under award numbers U01DA041048, U01DA050989, U01DA051016, U01DA041022, U01DA051018, U01DA051037, U01DA050987, U01DA041174, U01DA041106, U01DA041117, U01DA041028, U01DA041134, U01DA050988, U01DA051039, U01DA041156, U01DA041025, U01DA041120, U01DA051038, U01DA041148, U01DA041093, U01DA041089, U24DA041123, U24DA041147. A full list of supporters is available at https://abcdstudy.org/about/federal-partners/. A listing of participating sites and a complete listing of the study investigators can be found at https://abcdstudy.org/wp-content/uploads/2019/04/Consortium_Members.pdf. ABCD consortium investigators designed and implemented the study and/or provided data but did not necessarily participate in the analysis or writing of this report. This manuscript reflects the views of the authors and may not reflect the opinions or views of the NIH or ABCD consortium investigators. The ABCD data repository grows and changes over time. The ABCD data used in this report came from release 5.1, NIMH Data Archive Digital Object Identifier <u>10.15154/z563-zd24</u>.

The manuscript’s language style was enhanced through the utilization of artificial intelligence (ChatGTP). Figures were created with BioRender.com.

## Notes

### Funding Statement

This study was funded by a fellowship from the University Medicine Essen Clinician Scientist Academy (UMEA) of the Medical Faculty of the University Duisburg-Essen, supported by the German Research Foundation (DFG), awarded to L.D. (FU 356/12-2).

### Author Declarations

The Institutional Review Board of the University of California, San Diego, gave ethical approval for the Adolescent Brain Cognitive Development (ABCD) study. The Ethics Committee of the Medical Faculty, University of Duisburg-Essen, gave ethical approval for the present analyses (24-12040-BO).

