## Supplemental Material for "The Sex-Specific Role of Adrenal Androgens in Youth Psychopathology"

### **Supplementary Methods – ABCD sample analyses**

Study Sample

Supplementary Figure S1. Participant Selection and Final Sample Composition.

Exposure: Hormone values

Supplementary Figure S2. Final Hormone Values Baseline and 1-Year Follow-Up by Sex.

**Supplementary Figure S3. Log-Transformed** Hormone **Values by Sex.**

Outcome: Child Behavior Checklist (CBCL)-Scores

Supplementary Fi1gure S4. Distribution of CBCL Category Scores.

Covariates and Missingness

Supplementary Figure S5. Directed Acyclic Graph to Identify a Minimal Adjustment Set.

Supplementary Table S1. Comparison of Included and Excluded Participants at 2-Year Follow-Up

Supplementary Table S2. Comparison of Included and Excluded Participants at 3-Year Follow-Up.

Supplementary Table S3. Comparison of Included and Excluded Participants at 4-Year Follow-Up.

Supplementary Table S4. Baseline Hormone and CBCL Values by Inclusion in 4-Year Follow-Up.

Supplementary Figure S6. Histogram of Imputed DHEA Values Below Lower Limit of Detection.

Supplementary Figure S7. Histogram of Final DHEA Values Including Imputed and Outlier Data.

Supplementary Figure S8. Histogram of Imputed Estradiol Values Below Lower Limit of Detection.

Supplementary Figure S9. Histogram of Final Estradiol Values Including Imputed and Outlier Data.

Supplementary Figure S10. Histogram of Imputed Testosterone Value Below Lower Limit of Detection.

Supplementary Figure S11. Histogram of Final Testosterone Value Including Imputed and Outlier Data.

Linear Mixed Models - Assumptions

Supplementary Table S5. Testing Multicollinearity in Males in DHEA at 3-Year Follow-Up.

Supplementary Table S6. Testing Multicollinearity in Females in DHEA at 3-Year Follow-Up.

Supplementary Figure S12. QQ-Plots to Assess the Normal Distribution of Residuals in Males.

Supplementary Figure S13. QQ-Plots to Assess the Normal Distribution of Residuals in Females.

Supplementary Figure S14. Evaluating Non-Linear Effects in Crude and Adjusted Models by Sex.

Supplementary Table S7. GAMMs Evaluating Non-Linear Effects of DHEA at 3-Year Follow-Up.

Sensitivity Analysis

Supplementary Table S8. Sensitivity Analysis – Negative Binomial Mixed Models for Males.

Supplementary Table S9. Sensitivity Analysis – Negative Binomial Mixed Models for Females.

Supplementary Table S10. Sensitivity Analysis – Excluding Males on Medication at Baseline.

Supplementary Table S11. Sensitivity Analysis – Excluding Females Medication at Baseline.

Supplementary Table S12. Sensitivity Analysis – Adjusting for PDS Sum Score at Baseline in Males.

Supplementary Table S13. Sensitivity Analysis – Adjusting for PDS Sum Score at Baseline in Females.

Supplementary Table S14. Sensitivity Analysis – Adjusting for PDS Sum Score at 3-Year in Males.

Supplementary Table S15. Sensitivity Analysis – Adjusting for PDS Sum Score at 3-Year in Females.

Supplementary Table S16. Sensitivity Analysis – Excluding Males with Hormone Levels below LLD.

Supplementary Table S17. Sensitivity Analysis – Excluding Females with Hormone Levels below LLD.

Supplementary Figure S15. Sensitivity Analysis – Sex-Stratified Associations Between DHEA Levels and CBCL Scores Across Follow-Ups in Raw, Adjusted, and Baseline-CBCL-Adjusted Models

**Supplementary Results**

Supplementary Table S18. Correlations between Hormones across Timepoints.

Supplementary Figure S16. Associations of DHEA with DSM-5 Problem Scores at 2-Year Follow-Up.

Supplementary Figure S17. Associations of DHEA with DSM-5 Problem Scores at 3-Year Follow-Up.

Supplementary Figure S18. Associations of DHEA with DSM-5 Problem Scores at 4-Year Follow-Up.

Supplementary Figure S19. Sex-Stratified Risk Ratios for the Associations between DHEA Levels and Child Behavior Checklist (CBCL) Diagnostic Categories.

Supplementary Table S19. DHEA × Sex-Interaction Effects on CBCL Outcomes Across Follow-Ups.

Supplementary Methods – ABCD sample analyses

*Study Sample*

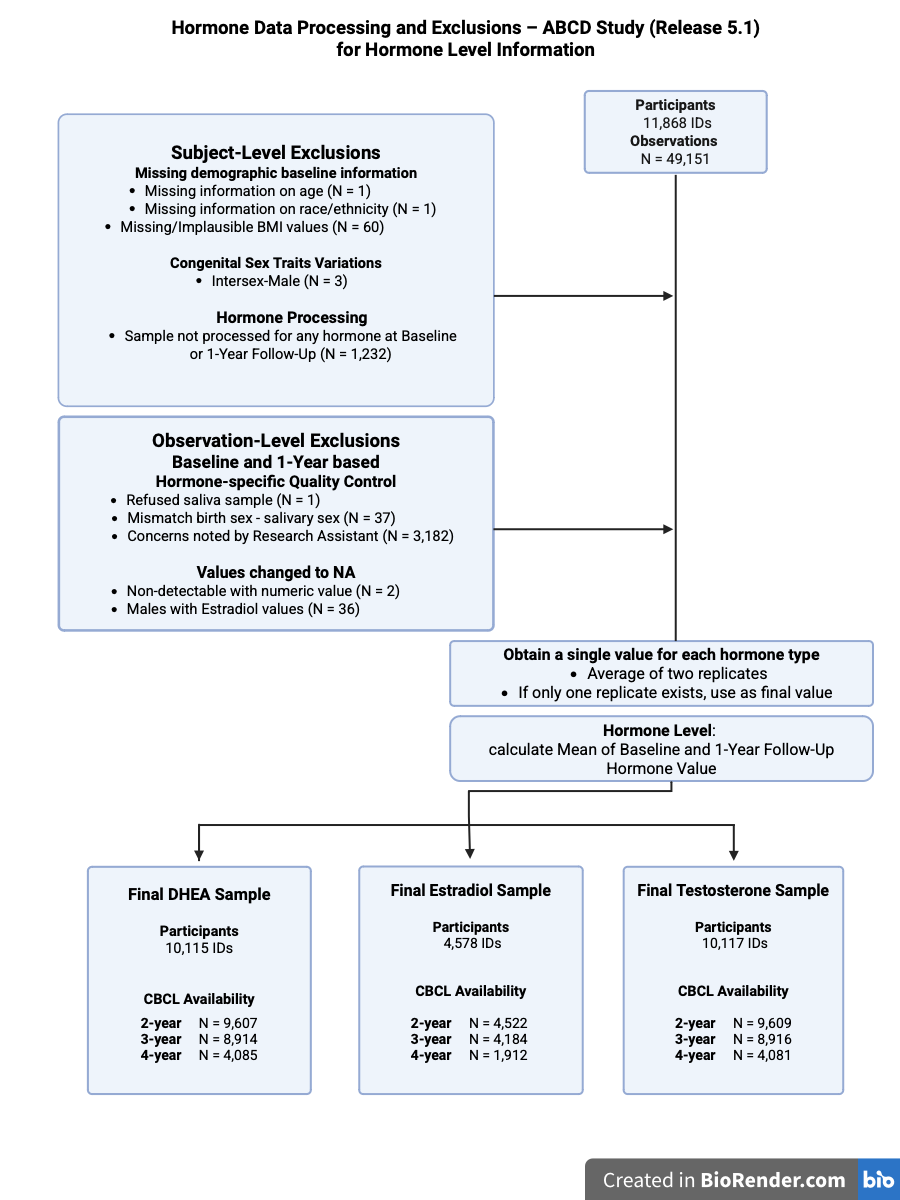

**Supplementary Figure S1. Participant Selection and Final Sample Composition.** This figure depicts the exclusion criteria applied to derive the final study sample used for the Hormone Level analyses, separately for each hormone and timepoint. Hormone Level is defined as the arithmetic mean of the log-transformed baseline and 1-year follow-up hormone values per participant. For example: DHEA Level = log(mean of baseline and 1-year follow-up DHEA value). If only baseline (N=1,052) or 1-year follow-up (N=1,465) was available, that single time point was used. Birth sex refers to the sex assigned at birth. Salivary sex indicates the sex recorded on the saliva sample tube. The variable ‘mismatch birth sex – salivary sex’ identifies participants for whom these two values differ.

The figure was created using Biorender.com.

*Exposure: Final Hormone Values Prior to Level and Change Calculations*

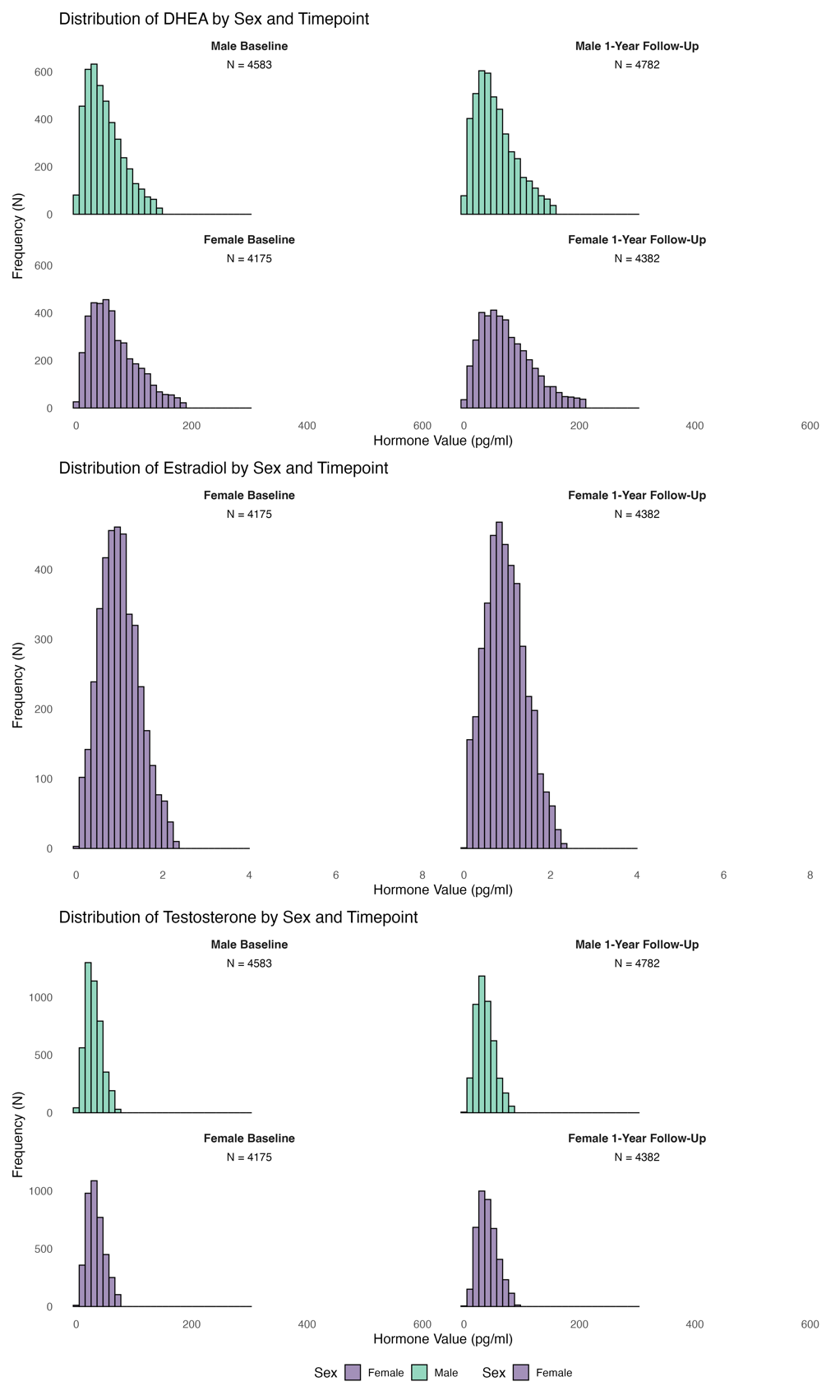

**Supplementary Figure S2. Hormone Values Baseline and 1-Year Follow-Up by Sex.** Final raw hormone values represent the average of two replicates per hormone and serve as the basis for calculating the hormone level and change variables. The x-axis limits of the histograms were adjusted to highlight the main distribution range of hormone values. Values outside these limits are not presented (N = 6). The observed outliers were as follows: DHEA: Male 1-Year Follow-Up: 615.70 ; Female Baseline: 648.62; Female 1-Year Follow-Up: 669.07, 933.42; Testosterone: Male 1-Year Follow-Up: 615.06; Female 1-Year Follow-Up: 1592.79 pg/ml.

*Exposure: Log-Transformed Hormone Levels and Changes*

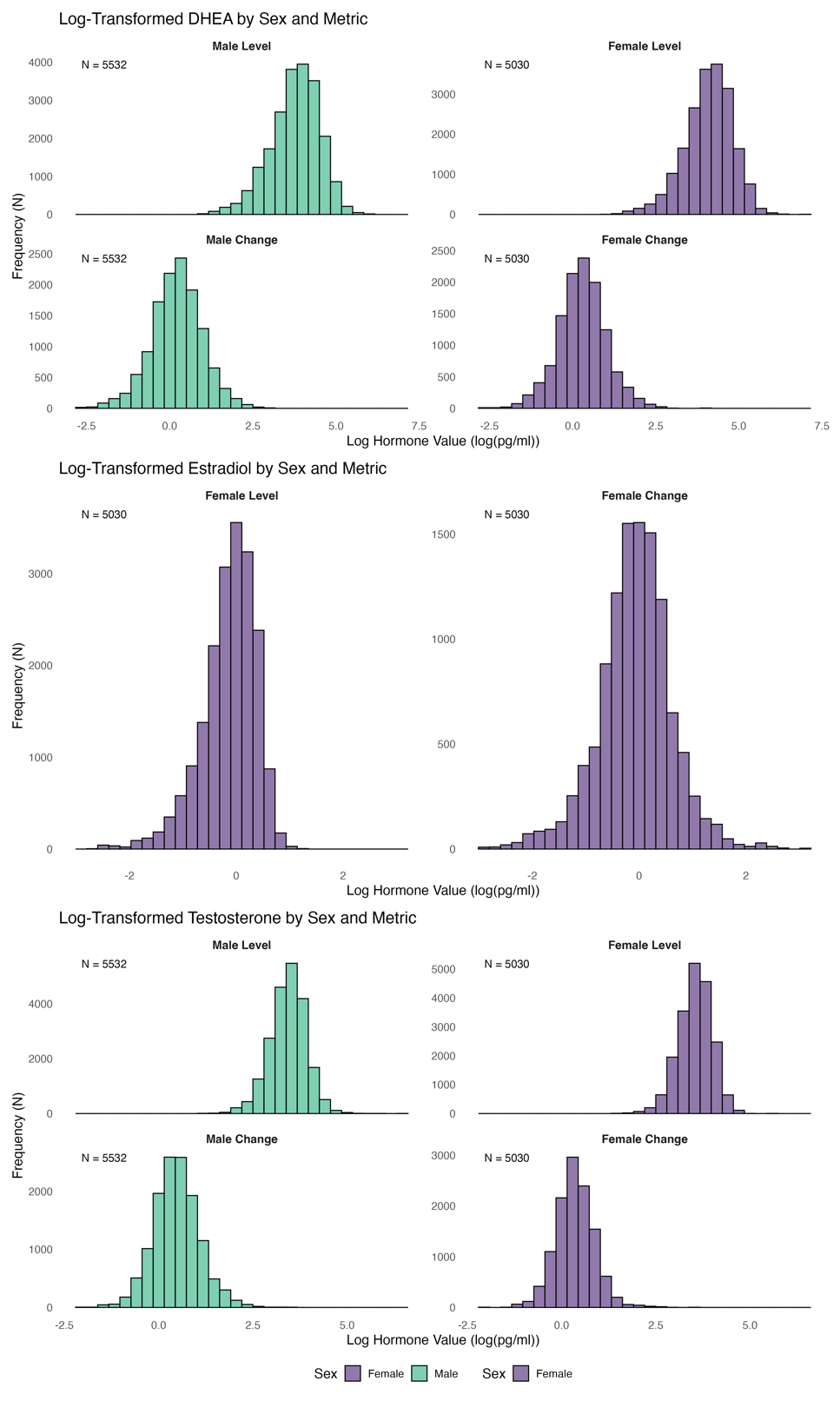

**Supplementary Figure S3. Log-Transformed Hormone Values by Sex.** Histograms of log-transformed hormone values for DHEA, Estradiol, and Testosterone in Males and Females, separated by Hormone Level and Change. Hormone Level is defined as the natural logarithm of the arithmetic mean of baseline and 1-year follow-up hormone values per participant. Hormone Change is defined as the natural logarithm of the ratio between 2-Year Follow-Up and baseline hormone values per participant. For example: DHEA Level = log(mean of baseline and 1-year follow-up DHEA value); DHEA Change = log(ratio of 2-year follow-up to baseline DHEA). Note: Estradiol was only measured in females. *Outcome: Child Behavior Checklist (CBCL)*

The Child Behavior Checklist (CBCL) is a standardized parent-report questionnaire assessing emotional and behavioral problems in children and adolescents over the past six months. It includes 113 items rated on a three-point Likert scale (0 = not true, 1 = somewhat true, 2 = very true). The CBCL provides symptom-oriented syndrome scales (e.g., Anxious/Depressed, Aggressive Behavior) and composite scores for Total Problems, Externalizing Problems, and Internalizing Problems, with higher scores indicating greater difficulties. In addition, DSM-5–oriented subscales are available, aligning items with clinical diagnostic categories such as ADHD and Anxiety Disorders.

For this study, primary analyses focused on the syndrome-oriented composite scales “CBCL Externalizing” (Rule-Breaking and Aggressive Behavior) and “CBCL Internalizing” (Anxious/Depressed, Withdrawn/Depressed, Somatic Complaints). The six individual DSM-5–oriented subscales were explored in secondary analyses. Importantly, raw scores (the sum of item ratings) were used instead of standardized T-scores, preserving the original metric and facilitating longitudinal modeling of symptom changes. An exception was the calculation of risk ratios, for which CBCL categories of normal, borderline, and clinical were based on standardized T-scores, as these clinical thresholds rely on T-score cutoffs. CBCL data from the 2-, 3-, and 4-year follow-ups served as outcome measures to assess associations with earlier hormonal profiles.

*Outcome: Child Behavior Checklist (CBCL) Scores – DHEA final values*

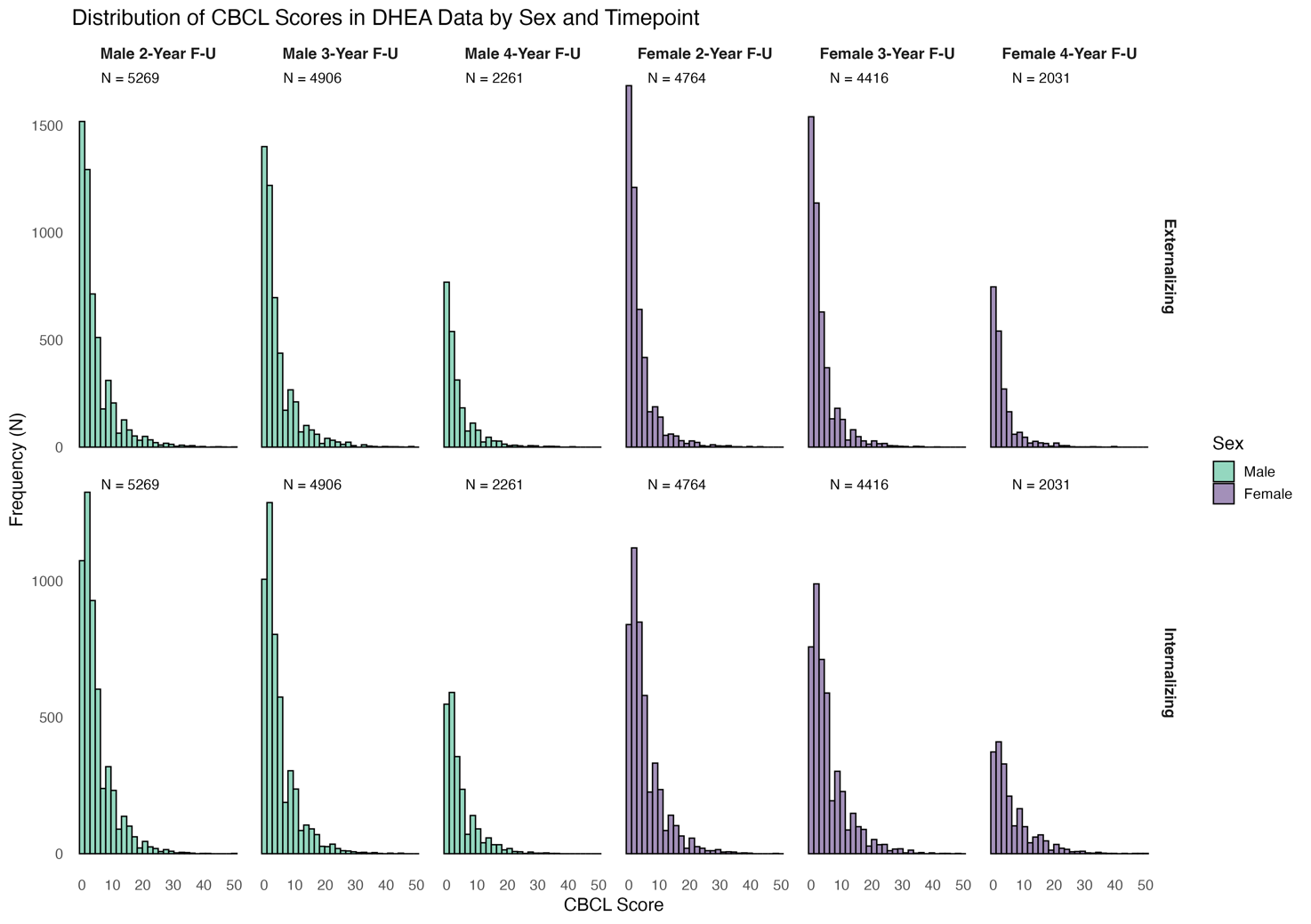
 **Supplementary Figure S4. Distribution of CBCL Category Scores in DHEA Sample by Sex.** Histograms showing the distribution of Externalizing and Internalizing CBCL scores across follow-up years (2-Year, 3-Year, and 4-Year Follow-Up) for male and female participants in the DHEA sample. Abbreviation: F-U = Follow-Up.

*Covariates – Directed Analytic Graph (DAG)*

Directed Acyclic Graphs (DAGs) are graphical representations of assumed causal relationships between variables. We used DAGs to identify potential confounders and covariates with DHEA as the primary hormone of interest. The identified adjustment set was then applied to models examining the effects of testosterone and estradiol as well. DAGs help to differentiate potential confounders from mediators, and colliders, and help to evaluate associations between confounders. Thus, they help to identify a minimally sufficient adjustment set for statistical analyses to minimize confounder bias.

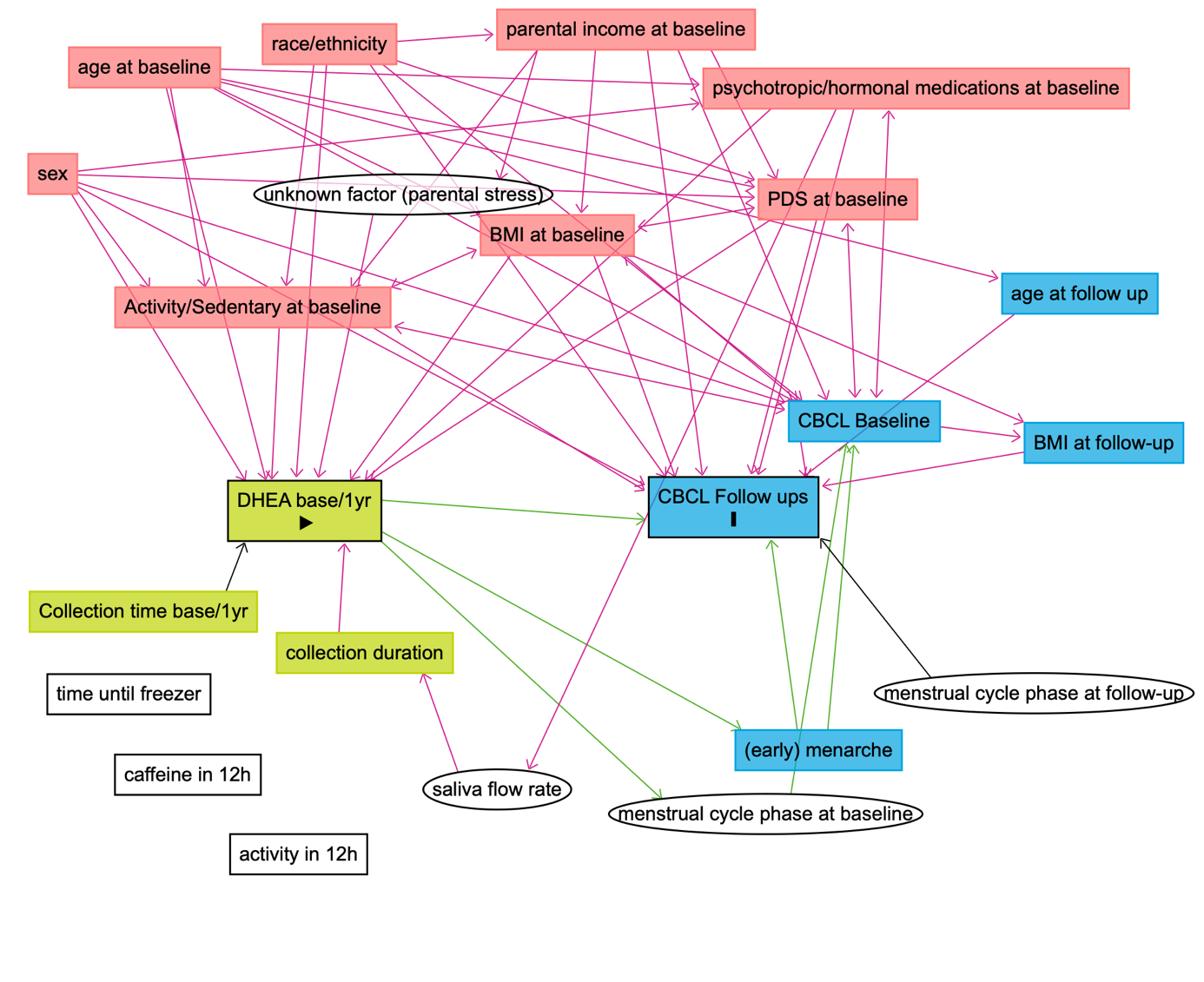

**Supplementary Figure S5. Directed Acyclic Graph to Identify a Minimal Adjustment Set.** Directed Acyclic Graph (DAG) illustrating assumed causal relationships between exposure (DHEA levels at baseline/1-year follow-up), outcome (CBCL outcomes at 2-, 3-, and 4-year follow-ups), and covariates. The DAG was created using DAGitty (www.dagitty.net;^1^). Light green boxes represent the exposure and its ancestors; blue boxes indicate the outcome and its ancestors. Red boxes denote shared ancestors of both exposure and outcome (i.e., potential confounders), while white boxes refer to unmeasured variables. Green arrows represent hypothesized causal paths; pink arrows indicate biasing paths (e.g., due to confounding). This visualization informed covariate selection for statistical adjustment in our models.

Based on theoretical reasoning and prior literature, causal directions in the DAG were specified as follows:

**Age at baseline:** DHEA levels begin to rise around age 6 in girls and 8 in boys and increase through puberty, indicating that age influences DHEA levels^2,3^. Age-related changes in psychopathology further justify age → CBCL^4^.

BMI SDS at baseline: Higher BMI is associated with increased adrenal androgens in prepubertal children^5^, supporting BMI → DHEA. Higher childhood BMI predicts internalizing problems such as anxiety and social withdrawal^6,7^. Bidirectional effects exist, but evidence supports BMI as an early factor^8,9^.

**Race/Ethnicity**: Ethnic differences in DHEA levels exist even pre-adrenarche^10^, and race/ethnicity also influences symptom expression and diagnosis in youth^11,12^.

**Physical activity/sedentary behavior**: Physical activity may alter DHEA^13,14^ and is associated with emotional wellbeing^15^. Activity itself is influenced by sex, age, BMI, CBCL, and socioeconomic status^16-18^.

**Pubertal development (PDS):** More advanced pubertal stage corresponds to higher DHEA levels^19,20^ and earlier timing predicts increased psychopathology^21,22^. Associations with BMI are also documented^23^. It has been proposed that controlling for pubertal stage should be avoided when examining the effects of pubertal hormones on mental health^24^, as hormones drive physical changes, making pubertal development a potential confounder along the causal pathway from hormone → pubertal development → mental health. This reasoning applies to gonadal hormones such as testosterone and estradiol, which directly underlie the physical changes of puberty^25^. However, it is less applicable to DHEA, which produces only subtle physical changes, making a causal pathway of pubertal development → DHEA → mental health more plausible (as shown). To remain consistent across evaluated hormones and to avoid the loss of participants due to missing PDS observations, we decided against controlling for PDS at baseline in the primary models but included a sensitivity analysis controlling for the PDS score at baseline as an additional covariate for the DHEA-models.

**Prescribed medication at baseline:** Antipsychotic medication is associated with elevated DHEA^26^. Psychopathology and medication have bidirectional influence, but CBCL scores typically precede pharmacological treatment. To assess effect modulation with medication use, a sensitivity analyses was conducted.

**Socioeconomic Status via Parental/Household income:** Lower household income is associated with reduced DHEA^27^ and increased behavioral/emotional problems^28,29^. However, a direct effect of household income on hormonal levels is physiologically implausible. Income also predicts physical activity, BMI and pubertal timing^20,30,31^, all with known associations to hormonal levels, rendering these variables plausible mediators of the income → DHEA relationship. Therefore, the minimally sufficient adjustment set included these variables but not the parental income.

**Baseline CBCL**: Baseline CBCL is a strong predictor of later CBCL scores, reflecting symptom continuity^32^. However, it remains unclear whether adrenal androgens influence psychopathological symptoms, implying that baseline CBCL scores might act as a mediator and should not be adjusted for, or whether the relationship operates in the opposite direction, making baseline CBCL scores a potential confounder (see Background section of the main manuscript). Given our primary hypothesis, namely that adrenal androgens influence youth psychopathology, we chose not to adjust for baseline CBCL scores in the main analyses. Nonetheless, we included them in extensive sensitivity analyses to assess the robustness of our findings.

Saliva Collection Context: Saliva collection time, caffeine intake, recent activity, and freezing delay were not found to systematically bias DHEA in previous studies^20,26,33^. Saliva flow rate may impact hormone concentration^34^ which was not recorded in our study. However, all these variables are unlikely to affect later psychopathology (measured via the CBCL) and therefore are unlikely to act as confounders in the current study.

Given these considerations, the following minimally sufficient adjustment set to estimate the total causal effect of DHEA at baseline on CBCL scores at later follow-ups was identified: baseline age (continuous), race/ethnicity (five-category factor), baseline BMI standard deviation score, and baseline physical activity (minutes per week). In separate sensitivity analyses, we included baseline psychopathology (CBCL scores), Pubertal Development Scale (PDS) sum score as additional covariates, and excluded participants with psychotropic or hormone-related medication use.

All models were estimated using linear mixed-effects models (LMMs) implemented in the *lme4* package in R. Besides the hormonal levels/changes and the above mentioned covariates as fixed factors, models additionally included a random intercept for study site (1 | site) and family nested within study site ((1 | site:family)) to account for clustering at both family and site levels. Separate models were run for each sex, hormone level and hormone change variable, and outcome.
Raw models included the exposure and Analyses were performed in R (version 4.4.3;^35^) using the lme4 package (version 1.1-37) for fitting linear mixed-effects models.

***Physical activity/sedentary behavior (SAIQ)***

Physical activity was operationalized using items from the ABCD Longitudinal Parent Sports and Activities Involvement Questionnaire (SAIQ) of the ABCD Study. For each reported sport, parents reported the number of days per week participants engaged in each activity as well as the average time spent per session. Because only physical activity and sedentary behavior at baseline may influence both hormone levels and mental health outcomes, we restricted analyses to baseline data.

Following the ABCD scoring protocol, frequency of participation was assessed on a categorical scale (1–7 days per week, once every two weeks, once a month, less than once a month). The response option “Don’t know” was coded as missing. Average duration per session was reported in minutes, with response options ranging from <30 minutes to >180 minutes. For analysis, <30 minutes was recoded as 15 minutes and >180 minutes as 210 minutes; “Don’t know” was again treated as missing.

Weekly minutes of activity were computed as the product of days per week and minutes per session. Total weekly physical activity was then derived as the sum across all reported sports and activities.

The following activities were included: ballet/dance, baseball/softball, basketball, climbing, field hockey, football, gymnastics, ice hockey, horseback riding/polo, ice or inline skating, martial arts, lacrosse, rugby, skateboarding, skiing/snowboarding, soccer, surfing, swimming/water polo, tennis, track/running, mixed martial arts (MMA), volleyball, yoga.

Activities not included due to their minimal contribution to physical exertion were: (1) playing a musical instrument, (2) visual arts (drawing, painting, graphic art, photography, pottery), (3) drama, theater, acting, film drama, (4) crafts (e.g., knitting, building model cars or airplanes), (5) competitive games (e.g., chess, cards, darts), and (6) hobbies such as collecting stamps or coins.

The resulting variable reflects total minutes of sports-related physical activity per week, which was included as covariate in all adjusted models.

*Missingness*

Initially, there were 49,151 observations from 11,868 participants. After applying exclusion criteria, 41,910 observations (85.27%) from 10,562 participants (89.00%) remained across all time points and hormones.

At the subject level, we excluded participants with missing values on key demographic variables (age at interview, race/ethnicity, missing or implausible BMI values), as well as those whose saliva samples were not processed for any hormone at either the baseline or 1-year follow-up time points. Additionally, participants with congenital differences in sex development were excluded.

At the observational level, we did not exclude entire participants but removed specific observations as part of hormone-specific quality control. Observations were excluded if the participant refused to provide a saliva sample, if there was a sex mismatch between sex assigned at birth and sex specified for the saliva sample, or if concerns were noted by the research assistant. Measurements labeled as “none [no hormone] detectable” that nonetheless produced values, as well as estradiol values for male participants, were set to missing values. Numbers are provided in Supplementary Figure 1.

For each follow-up year (2-, 3-, and 4-year), we generated a binary exclusion variable to identify observations meeting any of the exclusion criteria. We then compared excluded and included observations with respect to age, sex, race/ethnicity, and family income. Across all follow-up years, participants with excluded observations did not differ in age compared to participants with included observations but were less likely to be of white ethnicity and more likely to be of black or hispanic backgrounds. Additionally, excluded participants more frequently came from lower-income families (see Supplementary Tables S1–S3).

| **Supplementary Table S1. Comparison of Included and Excluded Participants at 2-Year Follow-Up.** | | | | |
| --- | --- | --- | --- | --- |
| **Characteristics** | | **Included**  **(N = 10,114)** | **Excluded**  **(N = 858)** | **p values ^a^**  **t-test/Χ²-test** |
| **Age (Years), mean (SD)** | | 12.03 (0.67) | 12.05 (0.65) | 0.389 |
| **Sex, N (%)** | |  |  |  |
|  | **Male** | 5,312 (52.52) | 441 (51.34) | < 0.001 |
|  | **Female** | 4,802 (47.48) | 415 (48.31) |  |
|  | **Intersex-Male** | 0 (0.00) | 3 (0.35) |  |
| **Race/Ethnicity, N (%)** | |  |  |  |
|  | White | 5,501 (54.39) | 360 (41.91) | < 0.001 |
|  | Black | 1,368 (13.53) | 193 (22.48) |  |
|  | Hispanic | 1,965 (19.43) | 199 (23.17) |  |
|  | Asian | 223 (2.20) | 8 (0.93) |  |
|  | Other/Multiethnic | 1,057 (10.45) | 99 (11.53) |  |
| **Family Income (past year) ^a^, N (%)** | |  |  |  |
|  | < $5000 | 347 (3.43) | 56 (6.52) | < 0.001 |
|  | $5,000 - $11,999 | 364 (3.60) | 59 (6.87) |  |
|  | $12,000 - $15,999 | 231 (2.28) | 37 (4.31) |  |
|  | $16,000 - $24,999 | 481 (4.76) | 40 (4.66) |  |
|  | $25,000 - $34,999 | 609 (6.02) | 68 (7.91) |  |
|  | $35,000 - $49,999 | 872 (8.62) | 76 (8.85) |  |
|  | $50,000 - $74,999 | 1,378 (13.62) | 121 (14.09) |  |
|  | $75,000 - $99,999 | 1,504 (14.87) | 108 (12.57) |  |
|  | $100,000 - $199,999 | 3,141 (31.06) | 230 (26.78) |  |
|  | > $200,000 | 1,187 (11.74) | 64 (7.45) |  |

Abbreviations: N = Number, SD = Standard deviation.

a. For categorical data (sex, race/ethnicity, family income class), Chi-squared tests were calculated. For numeric data (age), a t-test was calculated.

| **Supplementary Table S2. Comparison of Included and Excluded Participants at 3-Year Follow-Up.** | | | | |
| --- | --- | --- | --- | --- |
| **Characteristics** | | **Included**  **(N = 9,530)** | **Excluded**  **(N = 805)** | **p values ^a^**  **t-test/Χ²-test** |
| **Age (Years), mean (SD)** | | 12.91 (0.65) | 12.92 (0.63) | 0.633 |
| **Sex, N (%)** | |  |  |  |
|  | **Male** | 5,001 (52.48) | 423 (52.48) | < 0.001 |
|  | **Female** | 4,529 (47.52) | 380 (47.15) |  |
|  | **Intersex-Male** | 0 (0.00) | 3 (0.37) |  |
| **Race/Ethnicity, N (%)** | |  |  |  |
|  | White | 5,237 (54.96) | 355 (44.10) | < 0.001 |
|  | Black | 1,198 (12.57) | 163 (20.25) |  |
|  | Hispanic | 1,873 (19.66) | 202 (25.10) |  |
|  | Asian | 214 (2.25) | 7 (0.87) |  |
|  | Other/Multiethnic | 1,008 (10.58) | 78 (9.69) |  |
| **Family Income (past year) ^a^, N (%)** | |  |  |  |
|  | < $5000 | 302 (3.17) | 43 (5.33) | < 0.001 |
|  | $5,000 - $11,999 | 317 (3.33) | 46 (5.71) |  |
|  | $12,000 - $15,999 | 215 (2.26) | 36 (4.47) |  |
|  | $16,000 - $24,999 | 437 (4.59) | 43 (5.33) |  |
|  | $25,000 - $34,999 | 556 (5.83) | 61 (7.57) |  |
|  | $35,000 - $49,999 | 811 (8.51) | 76 (9.43) |  |
|  | $50,000 - $74,999 | 1,309 (13.74) | 107 (13.28) |  |
|  | $75,000 - $99,999 | 1,421 (14.91) | 104 (12.90) |  |
|  | $100,000 - $199,999 | 3,012 (31.61) | 221 (27.42) |  |
|  | > $200,000 | 1,150 (12.07) | 69 (8.56) |  |

Abbreviations: N = Number, SD = Standard deviation.

a. For categorical data (sex, race/ethnicity, family income class), Chi-squared tests were calculated. For numeric data (age), a t-test was calculated.

| **Supplementary Table S3. Comparison of Included and Excluded Participants at 4-Year Follow-Up.** | | | | |
| --- | --- | --- | --- | --- |
| **Characteristics** | | **Included**  **(N = 4,366)** | **Excluded**  **(N = 388)** | **p values ^a^**  **t-test/Χ²-test** |
| **Age (Years), mean (SD)** | | 14.08 (0.68) | 14.10 (0.66) | 0.465 |
| **Sex, N (%)** | |  |  |  |
|  | **Male** | 2,293 (52.52) | 195 (50.26) | 0.003 |
|  | **Female** | 2,073 (47.48) | 192 (49.48) |  |
|  | **Intersex-Male** | 0 (0.00) | 1 (0.26) |  |
| **Race/Ethnicity, N (%)** | |  |  |  |
|  | White | 2,497 (57.20) | 185 (47.68) | < 0.001 |
|  | Black | 444 (10.17) | 65 (16.75) |  |
|  | Hispanic | 882 (20.20) | 97 (25.00) |  |
|  | Asian | 107 (2.45) | 3 (0.77) |  |
|  | Other/Multiethnic | 436 (9.99) | 38 (9.79) |  |
| **Family Income (past year) ^a^, N (%)** | |  |  |  |
|  | < $5000 | 124 (2.84) | 21 (5.41) | < 0.001 |
|  | $5,000 - $11,999 | 134 (3.07) | 20 (5.15) |  |
|  | $12,000 - $15,999 | 90 (2.06) | 18 (4.64) |  |
|  | $16,000 - $24,999 | 187 (4.28) | 16 (4.12) |  |
|  | $25,000 - $34,999 | 264 (6.05) | 27 (6.96) |  |
|  | $35,000 - $49,999 | 383 (8.77) | 33 (8.50) |  |
|  | $50,000 - $74,999 | 607 (13.90) | 53 (13.66) |  |
|  | $75,000 - $99,999 | 677 (15.51) | 49 (12.63) |  |
|  | $100,000 - $199,999 | 1,371 (31.40) | 117 (30.15) |  |
|  | > $200,000 | 529 (12.12) | 34 (8.76) |  |

Abbreviations: N = Number, SD = Standard deviation.

a. For categorical data (sex, race/ethnicity, family income class), Chi-squared tests were calculated. For numeric data (age), a t-test was calculated.

*Comparison of Baseline Hormone Levels and CBCL Values by Inclusion in 4-Year Follow-Up.*

We compare baseline hormone levels of DHEA, testosterone, estradiol and CBCL externalizing and internalizing scores between participants who contributed data to the 4-year follow-up and those who did not. Inclusion in the 4-year follow-up was defined based on the availability of DHEA measurements and CBCL scores. While group differences reached statistical significance, the absolute differences were small. Missingness primarily reflects the limited availability of 4-year follow-up data in release 5.1, in which only about half of the cohort was documented. Moreover, the longitudinal trends in DHEA and testosterone effects between 2- and 4-year follow-ups were consistent in both direction and magnitude, despite opposite patterns of missingness at baseline. For CBCL differences were only present for externalizing scores, and our findings remained robust when controlling for baseline CBCL (Supplementary Figure S15). Together, these considerations indicate that, while missingness is not completely at random, it is unlikely to account for the observed associations at the 4-year follow-up.

**Supplementary Table S4. Baseline Hormone and CBCL Values by Inclusion in 4-Year Follow-Up.**

| **Baseline Variable** | **Included in 4-year FU** | | | **Missing in 4-year FU** | | | **p-value** |
| --- | --- | --- | --- | --- | --- | --- | --- |
|  | N | Mean | SD | N | Mean | SD |  |
| DHEA Level | 4,085 | 3.95 | 0.73 | 6,030 | 3.90 | 0.76 | <0.001 |
| Testosterone Level | 4,073 | 3.47 | 0.47 | 6,044 | 3.51 | 0.47 | <0.001 |
| Estradiol Level | 1,912 | -0.08 | 0.48 | 2,866 | -0.20 | 0.53 | <0.001 |
| CBCL Externalizing Score | 4,085 | 4.16 | 5.40 | 7,775 | 4.61 | 6.10 | <0.001 |
| CBCL Internalizing Score | 4,085 | 4.98 | 5.32 | 7,775 | 5.09 | 5.64 | 0.287 |

Abbreviations: CBCL = Child Behavior Checklist; DHEA = Dehydroepiandrosterone; FU = Follow Up; N = Number.

***Imputation of Hormone Values Below the Lower Limit of Detection (LLD)***

Hormone values below the Lower Limit of Detection (LLD; DHEA: 5 pg/ml; Estradiol: 0.1 pg/ml; Testosterone: 1 pg/ml) were imputed on the log-transformed scale using age and sex as predictors (except for Estradiol, measured only in females). Imputation was conducted using a truncated normal distribution on the log scale to ensure imputed values did not exceed the LLD and to maintain the statistical properties of the transformed data. Prior log-transformation was applied to approximate a normal distribution of hormone values, thereby justifying the use of normal distribution-based imputation methods. Imputations were performed separately for each hormone replicate and each time point to retain valuable data otherwise lost due to censoring. Each final hormone value was then calculated as the average of both hormone replicates. The total number of imputed values was: DHEA Replicate 1: N = 308 (0.90%), DHEA Replicate 2: N = 321 (0.94%), Estradiol Replicate 1: N = 116 (0.71%), Estradiol Replicate 2: N = 109 (0.67%), Testosterone Replicate 1: N = 1 (0.003%), Testosterone Replicate 2: N = 8 (0.024%). All imputed values remained below their respective detection limits.

For each hormone, two log-transformed histograms are shown: the first illustrates the distribution of values imputed for measurements below the LLD on a log-transformed scale, while the second displays the full distribution of all hormone values, including imputed values and identified outliers. These plots demonstrate that the imputed values align well with the expected continuation of a log-normal distribution of the values above the LLD, consistent with physiological expectations.

*DHEA*

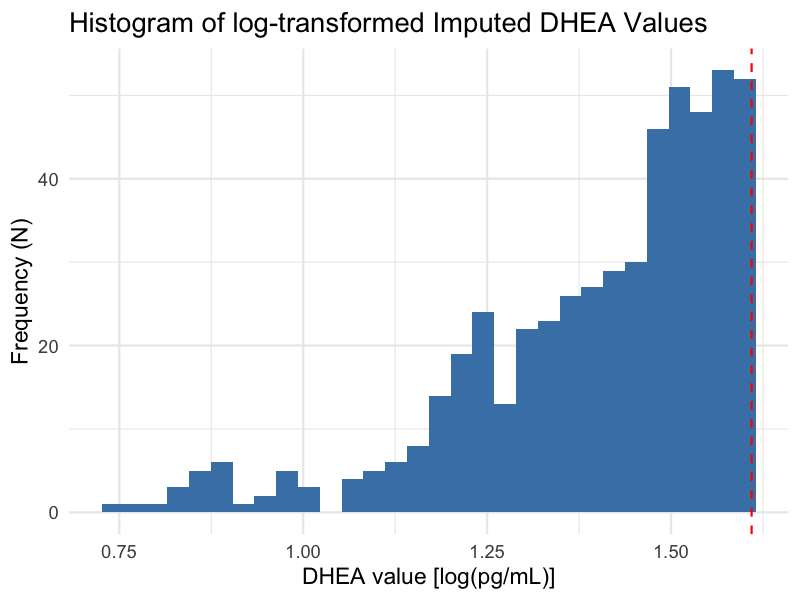

**Supplementary Figure S6. Histogram of Imputed DHEA Values Below Lower Limit of Detection.** The lower limit of detection (LLD) for DHEA is 5 pg/ml, corresponding to a log-transformed value (natural logarithm) of approximately 1.6.

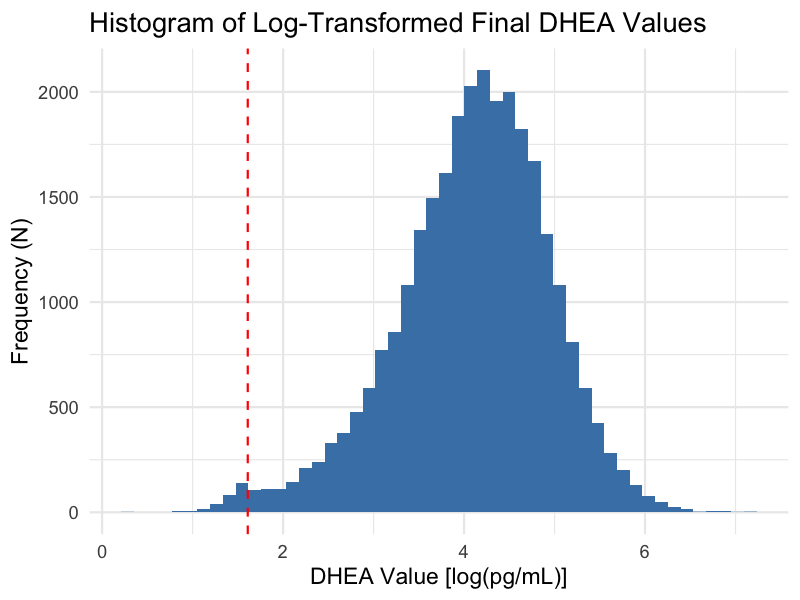

**Supplementary Figure S7. Histogram of Final DHEA Values Including Imputed and Outlier Data.**

*Estradiol*

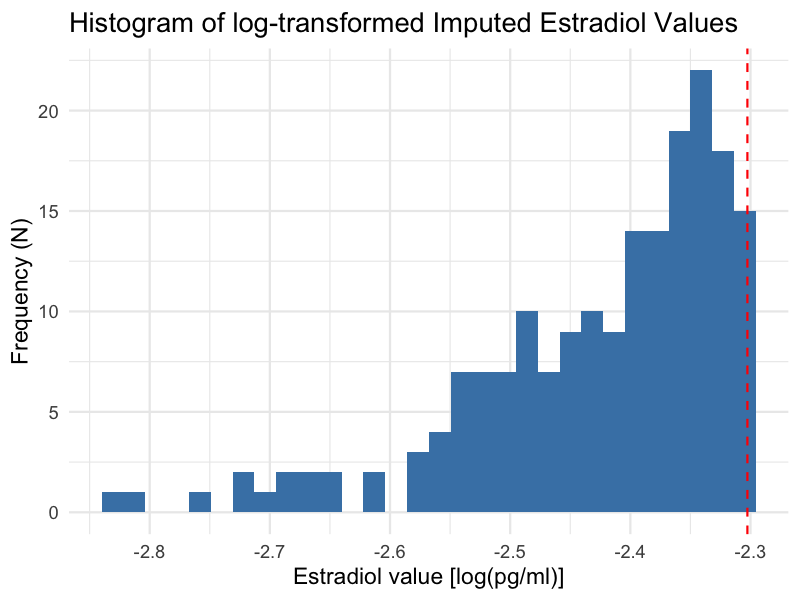

**Supplementary Figure S8. Histogram of Imputed Estradiol Values Below Lower Limit of Detection.** The lower limit of detection (LLD) for DHEA is 0.1 pg/ml, corresponding to a log-transformed value (natural logarithm) of approximately -2.3.

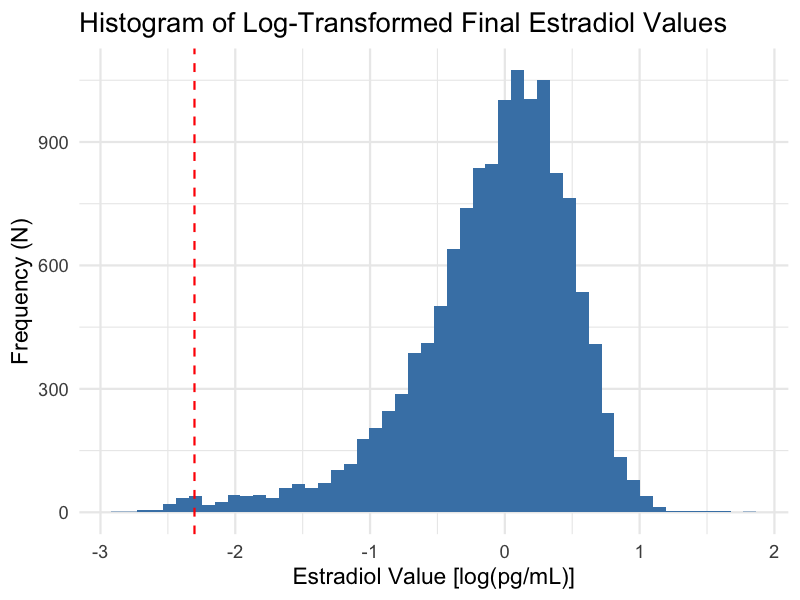

**Supplementary Figure S9. Histogram of Final Estradiol Values Including Imputed and Outlier Data.**

*Testosterone*

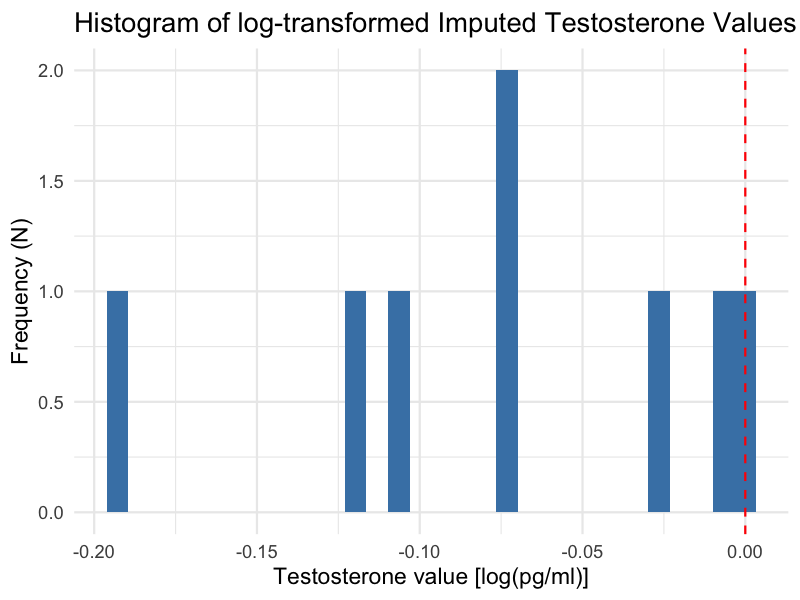

**Supplementary Figure S10. Histogram of Imputed Testosterone Value Below Lower Limit of Detection.** The lower limit of detection (LLD) for DHEA is 1 pg/ml, corresponding to a log-transformed value (natural logarithm) of 0.0.

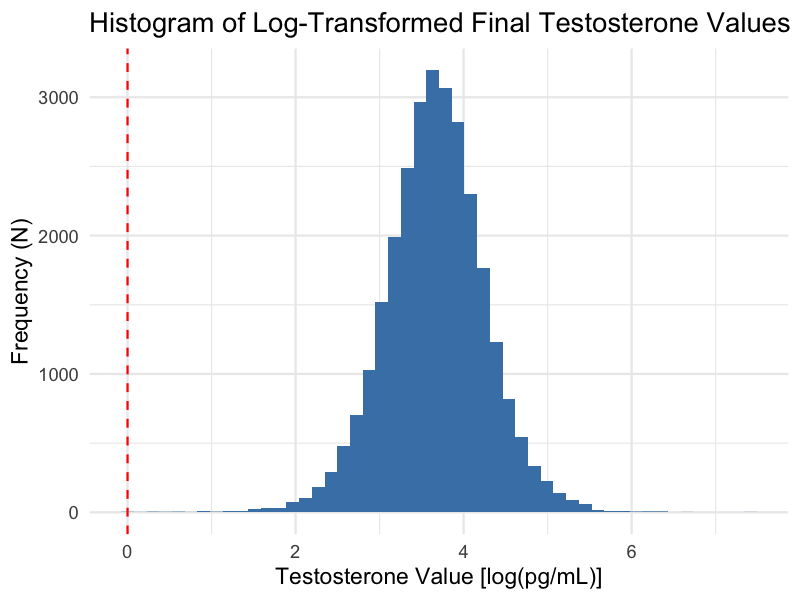

**Supplementary Figure S11. Histogram of Final Testosterone Value Including Imputed and Outlier Data.**

*Linear Mixed Models – Assumptions*

*Multicollinearity*

To assess multicollinearity among predictors, we computed generalized variance inflation factors (GVIFs) from linear models including only fixed effects (random effects were excluded). To enable comparisons across categorical variables with differing numbers of levels, GVIFs were scaled using the transformation GVIF^(1/(2*df)), and we interpreted the squared values of these adjusted GVIFs (i.e., GVIF^(1/df)). A squared adjusted GVIF exceeding a threshold of 4–5 was considered indicative of potentially problematic multicollinearity warranting further investigation. None of the covariates approached this threshold, indicating that problematic multicollinearity was unlikely.

**Supplementary Table S5. Testing Multicollinearity in Males in DHEA at 3-Year Follow-Up.**

|  | | **CBCL Externalizing** | | **CBCL Internalizing** | |
| --- | --- | --- | --- | --- | --- |
| **Variable** | | GVIF | GVIF^1/df^ | GVIF | GVIF^1/df^ |
|  | DHEA Level ^a^ | 1.11 | 1.12 | 1.11 | 1.11 |
|  | Age | 1.07 | 1.07 | 1.07 | 1.07 |
|  | Race/Ethnicity | 1.11 | 1.03 | 1.11 | 1.03 |
|  | BMI-SDS at baseline | 1.11 | 1.11 | 1.11 | 1.11 |
|  | Physical Activity | 1.04 | 1.03 | 1.04 | 1.04 |

Abbreviation: CBCL = Child Behavior Checklist, BMI-SDS = Body Mass Index – Standard Deviation Scores; GVIF = Generalized variance inflation factors. GVIF^1/df^ = squared adjusted GVIF.
^a^ DHEA Level = log(DHEA Mean of Baseline and 1-Year Follow-Up)

**Supplementary Table S6. Testing Multicollinearity in Females in DHEA at 3-Year Follow-Up.**

|  | | **CBCL Externalizing** | | **CBCL Internalizing** | |
| --- | --- | --- | --- | --- | --- |
| **Variable** | | GVIF | GVIF^1/df^ | GVIF | GVIF^1/df^ |
|  | DHEA Level ^a^ | 1.18 | 1.18 | 1.18 | 1.18 |
|  | Age | 1.10 | 1.10 | 1.10 | 1.10 |
|  | Race/Ethnicity | 1.13 | 1.03 | 1.13 | 1.03 |
|  | BMI-SDS at baseline | 1.13 | 1.13 | 1.13 | 1.13 |
|  | Physical Activity | 1.05 | 1.05 | 1.05 | 1.05 |

Abbreviation: CBCL = Child Behavior Checklist, BMI-SDS = Body Mass Index – Standard Deviation Scores; GVIF = Generalized variance inflation factors. GVIF^1/df^ = squared adjusted GVIF.
^a^ DHEA Level = log(DHEA Mean of Baseline and 1-Year Follow-Up)

*Normality of Residuals*

To assess the assumption of normally distributed residuals in the adjusted linear mixed-effects models, Quantile-Quantile (QQ) plots were generated for each outcome measure (CBCL Externalizing and CBCL Internalizing) at the 3-year follow-up, separately for males and females. These plots compare the quantiles of the standardized residuals to the theoretical quantiles of a normal distribution. Even though hormone levels were log-transformed prior to analysis to improve normality, visual inspection revealed deviations from normality, especially at the lower and upper extremes of the distribution. These presumably stem from the non-normal distribution of the outcome variable (see Supplementary Figure S4). Therefore, log-binomial distribution models were calculated as sensitivity analyses.

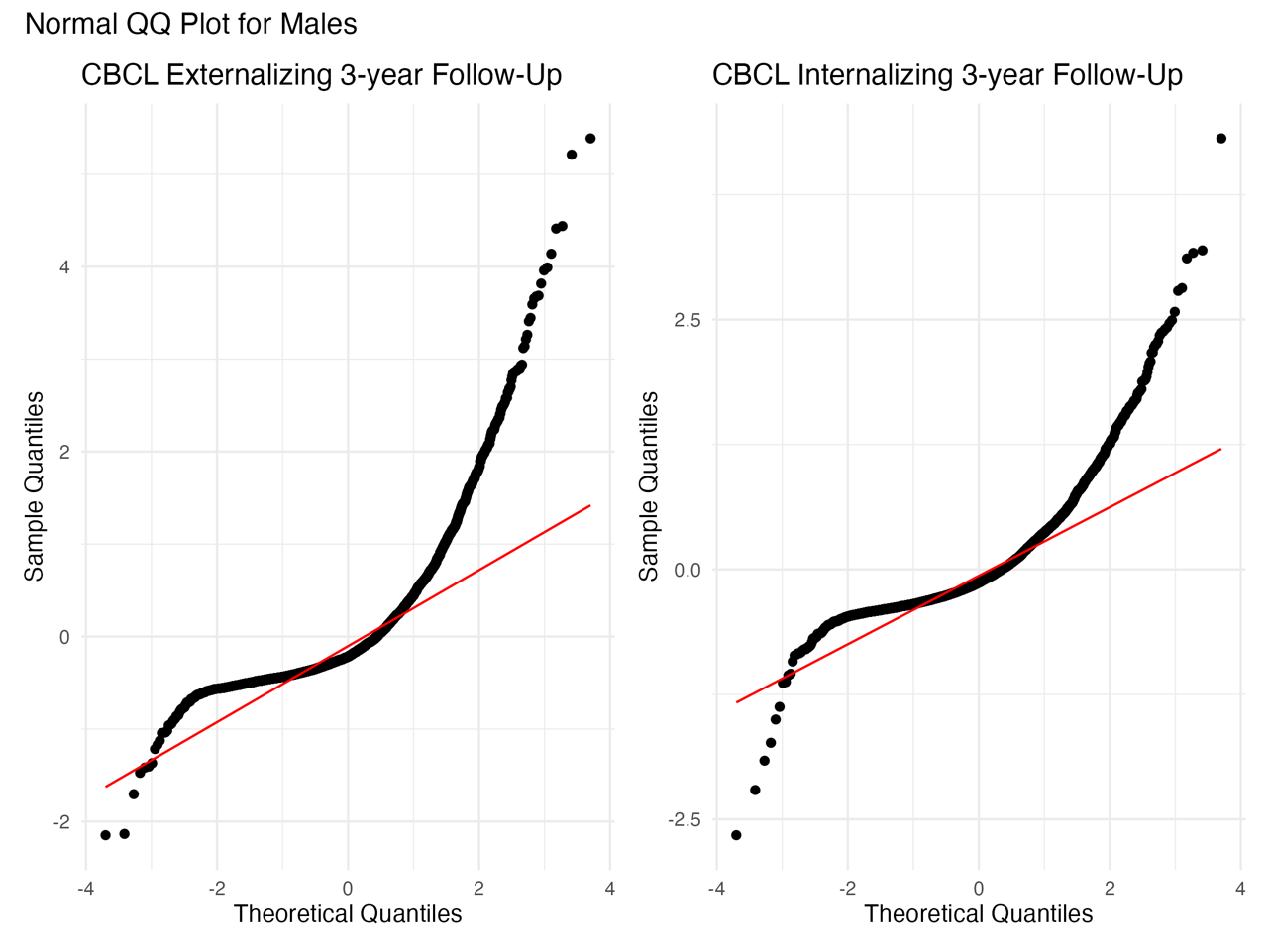

**Supplementary Figure S12. QQ-Plots to Assess the Normal Distribution of Residuals in Males.**

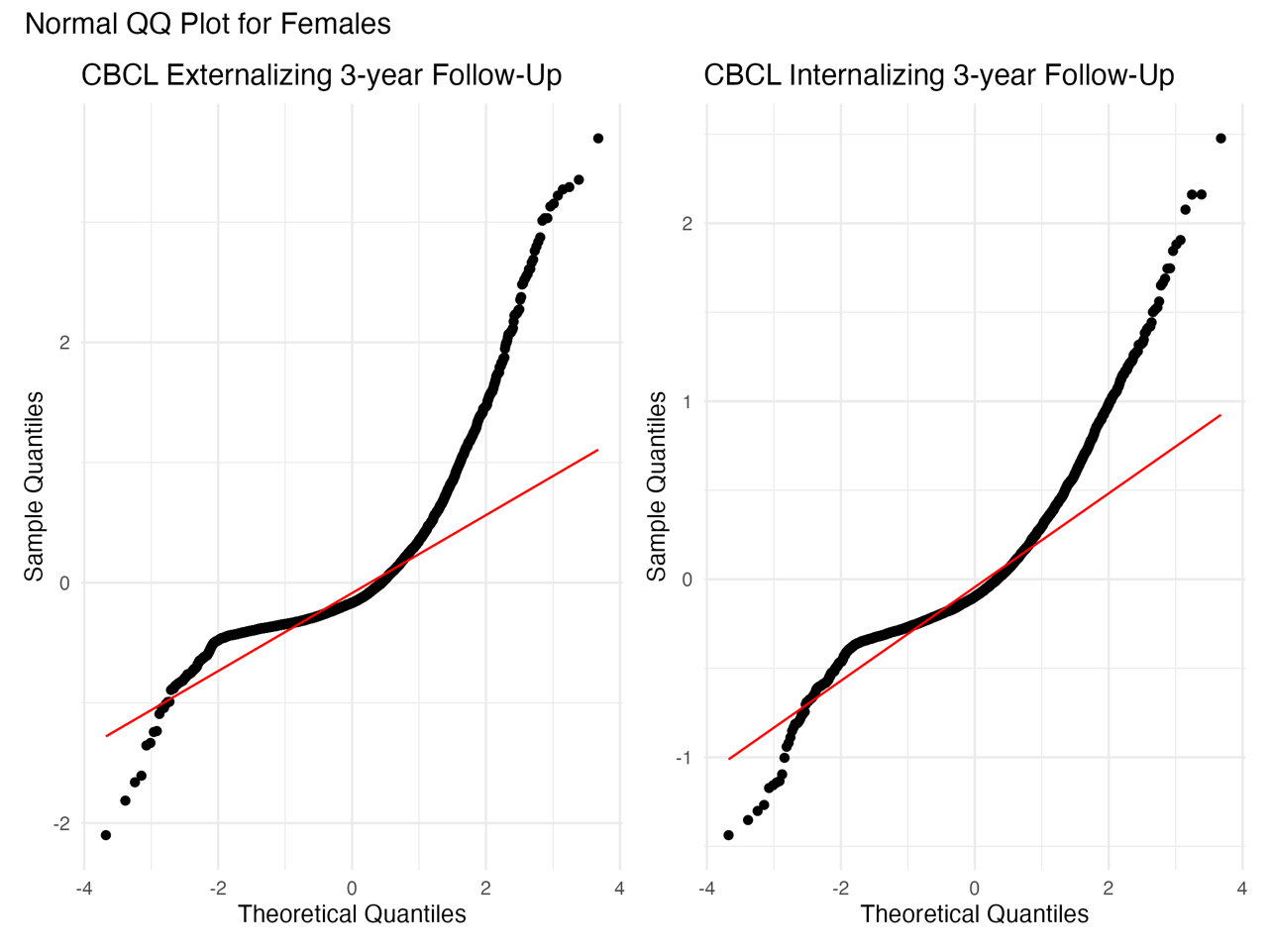

**Supplementary Figure S13. QQ-Plots to Assess the Normal Distribution of Residuals in Females.**

*Non-Linearity*

To evaluate the suitability of linear models, we first plotted the raw associations between hormone levels and CBCL outcomes to visually inspect potential non-linear relationships in the raw data (see Supplementary Figure S14), which compares raw and adjusted models to raw hormone values and indicates approximately linear associations. Additionally, we fitted Generalized Additive Mixed Models (GAMMs) incorporating penalized thin plate regression splines for DHEA level and DHEA change at the 2-year follow-up. These models were run separately for CBCL Externalizing and Internalizing symptoms, stratified by sex, and adjusted for age, race/ethnicity, BMI-SDS, Pubertal Development Score (PDS), and physical activity. Model comparisons using the Akaike Information Criterion (AIC), with a threshold of Δ AIC ≥ 2 indicating improved fit^36^, alongside likelihood ratio tests (LRT), showed no significant advantage of the more complex GAMMs over simpler linear models^37^. Given the visual evidence and these statistical results, as well as the greater interpretability of linear models – particularly important in clinical contexts – we focused our main analyses on linear effects.

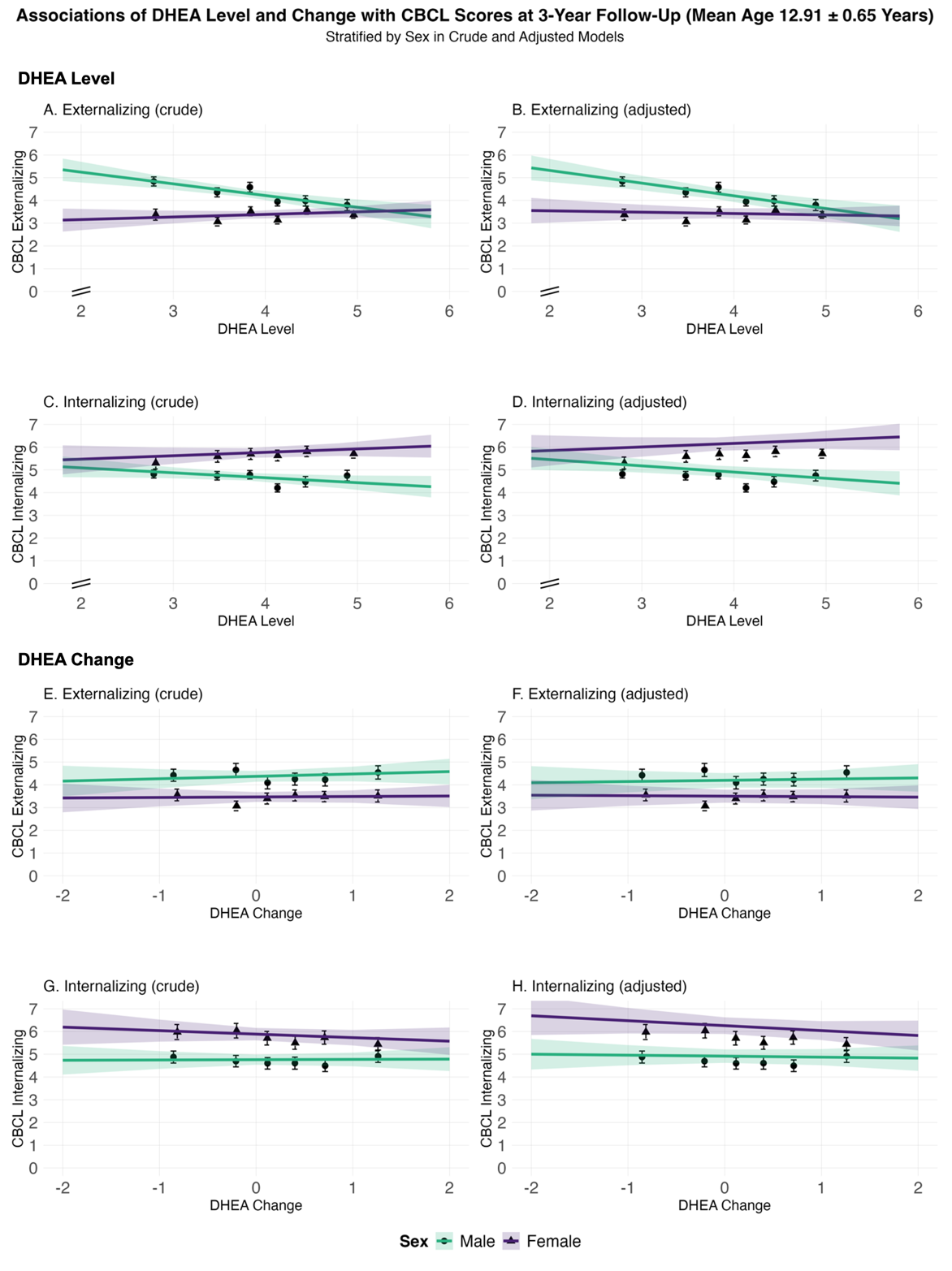

**Supplementary Figure S14. Evaluating Non-Linear Effects in Crude and Adjusted Models.** This figure displays sex-stratified associations between DHEA exposures and Child Behavior Checklist (CBCL) outcomes at 3-year follow-up. Separate panels show crude models (left column: A, C, E, G) and models adjusted for age, race/ethnicity, BMI SDS and physical activity assessed at baseline (right column: B, D, F, H), estimated using linear mixed-effect models.

DHEA exposures include DHEA Level (log-transformed mean of baseline and 1-year values; panels A–D) and DHEA Change (log-transformed ratio of 2-year to baseline; panels E–H). CBCL outcomes are Externalizing Problem Scores (A, B, E, F) and Internalizing Problem Scores (C, D, G, H). The x-axis for DHEA Level is truncated below x=2, as indicated by axis break. The y-axis represents CBCL scores (range: 0–7). Sample sizes: males (teal), N=5,001; females (purple), N=4,529. Error bars indicate 95% confidence intervals.

For complete data, including non-truncated plots, see Supplementary Figure S7.

Abbreviations: DHEA = dehydroepiandrosterone; CBCL = Child Behavior Checklist; BMI SDS = body mass index standard deviation score.

**Supplementary Table S7 – GAMM Results Evaluating Non-Linear Effects of DHEA at 3-Year Follow-Up.**

|  | | **Males** | | | | **Females** | | | |
| --- | --- | --- | --- | --- | --- | --- | --- | --- | --- |
| **Variable** | | EDF ^a^ | p_smooth_ ^b^ | Δ AIC ^c^ | p_LRT_ ^d^ | EDF ^a^ | p_smooth_ ^b^ | Δ AIC ^c^ | p_LRT_ ^d^ |
|  | Externalizing (Level, Raw) | 1.35 | <0.001 | -2.54 | 1.000 | 1.44 | 0.357 | -2.54 | 1.000 |
|  | Externalizing (Level, Adjusted) | 1.53 | <0.001 | -2.38 | 1.000 | 1.00 | 0.675 | -2.64 | 1.000 |
|  | Internalizing (Level, Raw) | 1.00 | 0.053 | -2.61 | 1.000 | 1.00 | 0.263 | -2.65 | 1.000 |
|  | Internalizing (Level, Adjusted) | 1.11 | 0.027 | -2.61 | 1.000 | 1.00 | 0.274 | -2.65 | 1.000 |
|  | Externalizing (Change, Raw) | 2.49 | 0.230 | -0.57 | 0.789 | 1.00 | 0.879 | -2.64 | 1.000 |
|  | Externalizing (Change, Adjusted) | 2.37 | 0.381 | -1.25 | 1.000 | 1.00 | 0.854 | -2.66 | 1.000 |
|  | Internalizing (Change, Raw) | 2.04 | 0.343 | -1.47 | 1.000 | 1.00 | 0.362 | -2.65 | 1.000 |
|  | Internalizing (Change, Adjusted) | 1.61 | 0.678 | -2.30 | 1. 000 | 1.00 | 0.208 | -2.66 | 1.000 |

Abbreviations: EDF = Effective Degrees of Freedom; GAMM = Generalized Additive Mixed Models; AIC = Akaike Information Criterion; LRT = Likelihood Ratio Test.

a. An EDF value of 1.00 indicates that the GAMM (Generalized Additive Mixed Model) estimated a linear relationship for DHEA Level or DHEA Change. Values of EDF greater than 1.00 indicate a non-linear (smoothed) term.
b. P-values representing the effect of DHEA Level or DHEA Change on Externalizing or Internalizing Symptoms, as estimated by a GAMM with a smoothed term (k = 5) for puberty timing (at baseline). Models included fixed effects for age, race/ethnicity, BMI-SDS, Pubertal Development Score (PDS), and physical activity. Random intercepts accounted for family nested within study site.
c. AIC difference between GAMMs with a smoothed versus a linear term for DHEA Level or DHEA Change. A negative Δ AIC indicates a better model fit with a linear term, a positive Δ AIC a better fit with a smoothed term; differences ≥ 2 were interpreted as relevant^36^.
d. P-values of the likelihood ratio test (LRT) evaluating whether the more complex model with a smoothed term significantly improves fit compared to the simpler linear model. P-values < 0.05 were considered statistically significant.

*Sensitivity Analyses – Negative binomial mixed models instead of linear mixed-effects models*

The outcome variables (CBCL Externalizing and Internalizing Scores) were highly right-skewed, with a large proportion of participants exhibiting low scores and only a small number showing very high values. To account for this distributional characteristic, we conducted a sensitivity analysis using negative binomial mixed models instead of linear mixed-effects models. Negative binomial models are specifically suited for overdispersed count-like or skewed continuous data, as they allow the variance to exceed the mean. By modeling the outcome with a log link function and an additional dispersion parameter, these models provide a more flexible framework for handling non-normally distributed residuals, thereby improving the robustness of effect estimates under distributional violations. Because the negative binomial family does not permit negative values, outcome variables were left unstandardized and only predictors were scaled, while standardized coefficients were obtained post hoc for comparability. The results from these models were largely consistent with those obtained from the primary analyses.

**Supplementary Table S8. Sensitivity Analysis – Negative Binomial Mixed Models for Males: Association of DHEA Levels at 3-Year Follow-Up.**

| **Outcome: CBCL Categories (Metric, Model)** |  | **Linear Mixed-Effects Models** | | | | **Negative Binominal Mixed Models** | | | |
| --- | --- | --- | --- | --- | --- | --- | --- | --- | --- |
|  |  | N_obs_ | b | 95%-CI | p | N_obs_ | b | 95%-CI | p |
| Externalizing (Level, Crude) |  | 4,676 | -0.06 | (-0.09, -0.03) | <0.001 | 4,676 | -0.09 | (-0.14, -0.05) | <0.001 |
| Externalizing (Level, Adjusted) |  | 4,676 | -0.07 | (-0.10, -0.04) | <0.001 | 4,367 | -0.10 | (-0.15, -0.06) | <0.001 |
| Internalizing (Level, Crude) |  | 4,676 | -0.03 | (-0.06, 0.00) | 0.064 | 4,676 | -0.03 | (-0.07, 0.01) | 0.108 |
| Internalizing (Level, Adjusted) |  | 4,676 | -0.03 | (-0.06, 0.00) | 0.022 | 4,367 | -0.04 | (-0.07, 0.00) | 0.053 |
| Externalizing (Change, Crude) |  | 2,733 | 0.01 | (-0.02, 0.05) | 0.453 | 2,733 | 0.00 | (-0.05, 0.06) | 0.873 |
| Externalizing (Change, Adjusted) |  | 2,733 | 0.01 | (-0.02, 0.05) | 0.500 | 2,573 | -0.00 | (-0.05, 0.05) | 0.986 |
| Internalizing (Change, Crude) |  | 2,733 | 0.00 | (-0.04, 0.04) | 0.943 | 2,744 | -0.01 | (-0.05, 0.04) | 0.820 |
| Internalizing (Change, Adjusted) |  | 2,733 | 0.00 | (-0.04, 0.04) | 0.962 | 2,573 | -0.01 | (-0.05, 0.03) | 0.660 |

Abbreviations: CBCL = Child Behavior Checklist, N_obs_ = Number of observations

**Supplementary Table S9. Sensitivity Analysis – Negative Binomial Mixed Models for Females: Association of DHEA Levels at 3-Year Follow-Up.**

| **Outcome: CBCL Categories (Metric, Model)** |  | **Linear Mixed-Effects Models** | | | | | | **Negative Binominal Mixed Models** | | | |
| --- | --- | --- | --- | --- | --- | --- | --- | --- | --- | --- | --- |
|  |  | N_obs_ | | b | | 95%-CI | p | N_obs_ | b | 95%-CI | p |
| Externalizing (Level, Crude) |  | 4,238 | 0.02 | | (-0.01, 0.05) | | 0.290 | 4,238 | 0.03 | (-0.02, 0.08) | 0.192 |
| Externalizing (Level, Adjusted) |  | 4,236 | 0.00 | | (-0.03, 0.03) | | 0.939 | 4,236 | 0.00 | (-0.05, 0.05) | 0.870 |
| Internalizing (Level, Crude) |  | 4,238 | 0.02 | | (-0.01, 0.05) | | 0.130 | 4,238 | 0.02 | (-0.02, 0.05) | 0.409 |
| Internalizing (Level, Adjusted) |  | 4,236 | 0.03 | | (0.00, 0.06) | | 0.084 | 4,236 | 0.02 | (-0.01, 0.06) | 0.221 |
| Externalizing (Change, Crude) |  | 2,495 | 0.00 | | (-0.04, 0.04) | | 0.919 | 2,495 | 0.00 | (-0.06, 0.06) | 0.935 |
| Externalizing (Change, Adjusted) |  | 2,494 | 0.00 | | (-0.04, 0.04) | | 0.916 | 2,494 | 0.00 | (-0.06, 0.06) | 0.936 |
| Internalizing (Change, Crude) |  | 2,495 | -0.02 | | (-0.05, 0.02) | | 0.362 | 2,495 | -0.01 | (-0.06, 0.03) | 0.530 |
| Internalizing (Change, Adjusted) |  | 2,494 | -0.02 | | (-0.06, 0.02) | | 0.261 | 2,494 | -0.02 | (-0.06, 0.02) | 0.385 |

Abbreviations: CBCL = Child Behavior Checklist, N_obs_ = Number of observations

*Sensitivity Analyses – Exclusion of Participants on Psychotropic or Hormone-Related Medication at Baseline*

Psychotropic or hormone-related medications may be associated with either the exposure (hormone levels and changes) or the outcome (CBCL scores). In the ABCD Study protocol, parents were asked to report any medications their child had taken during the past two weeks, with up to 15 entries possible. Reported medication names correspond to variants listed in the BioPortal RxNorm database (https://bioportal.bioontology.org). We extracted all medication entries across the 15 columns, yielding N = 973 unique medication names. From these, we manually identified all psychotropic and hormone-related medications, resulting in N = 291 distinct medication names. Those medications included stimulants (e.g., methylphenidate, amphetamines), antidepressants (e.g., fluoxetine, escitalopram), antipsychotics, and glucocorticoids. The three most frequently reported excluded medications were Albuterol (142 mentions), Methylphenidate (127 mentions), and Adderall (118 mentions). A complete list of 291 psychotropic and hormone-related medications is available from the authors upon request.

In total, 10.42% of the final study sample (N = 1,101) were reported by their parents to take such medications at baseline. Thus, we conducted a sensitivity analysis excluding these participants. Of these, N = 756 were males (13.67% of male participants) and N = 345 were females (6.86% of female participants). We then compared the results of these models to those from the primary analyses including all participants. The findings were consistent with the primary analyses.

**Supplementary Table S10. Sensitivity Analysis – Excluding Males on Medication at Baseline: Association of DHEA Levels and CBCL at 3-Year Follow-Up.**

| **Outcome: CBCL Categories (Metric, Model)** |  | **Primary Analysis: Participants with medication included** | | | | | **Sensitivity Analysis: Participants with medication excluded** | | | |
| --- | --- | --- | --- | --- | --- | --- | --- | --- | --- | --- |
|  |  | N_obs_ | b | | 95%-CI | p | N_obs_ | b | 95%-CI | p |
| Externalizing (Level, Crude) |  | 4,676 | -0.06 | (-0.09, -0.03) | | <0.001 | 4,058 | -0.06 | (-0.09, -0.03) | <0.001 |
| Externalizing (Level, Adjusted) |  | 4,676 | -0.07 | (-0.10, -0.04) | | <0.001 | 4,058 | -0.08 | (-0.11, -0.04) | <0.001 |
| Internalizing (Level, Crude) |  | 4,676 | -0.03 | (-0.06, 0.00) | | 0.064 | 4,058 | -0.02 | (-0.05, 0.01) | 0.210 |
| Internalizing (Level, Adjusted) |  | 4,676 | -0.03 | (-0.06, 0.00) | | 0.022 | 4,058 | -0.03 | (-0.06, 0.00) | 0.056 |
| Externalizing (Change, Crude) |  | 2,733 | 0.01 | (-0.02, 0.05) | | 0.453 | 2,378 | 0.00 | (-0.04, 0.05) | 0.825 |
| Externalizing (Change, Adjusted) |  | 2,733 | 0.01 | (-0.02, 0.05) | | 0.500 | 2,378 | 0.00 | (-0.04, 0.04) | 0.956 |
| Internalizing (Change, Crude) |  | 2,733 | 0.00 | (-0.04, 0.04) | | 0.943 | 2,378 | -0.01 | (-0.05, 0.03) | 0.616 |
| Internalizing (Change, Adjusted) |  | 2,733 | 0.00 | (-0.04, 0.04) | | 0.962 | 2,378 | -0.01 | (-0.05, 0.03) | 0.527 |

Abbreviations: CBCL = Child Behavior Checklist, N_obs_ = Number of observations

**Supplementary Table S11. Sensitivity Analysis – Excluding Females on Medication at Baseline: Association of DHEA Levels and CBCL at 2-Year Follow-Up.**

| **Outcome: CBCL Categories (Metric, Model)** |  | **Primary Analysis: Participants with medication included** | | | | **Sensitivity Analysis: Participants with medication excluded** | | | |
| --- | --- | --- | --- | --- | --- | --- | --- | --- | --- |
|  |  | N_obs_ | b | 95%-CI | p | N_obs_ | b | 95%-CI | p |
| Externalizing (Level, Crude) |  | 4,238 | 0.02 | (-0.01, 0.05) | 0.290 | 3,950 | 0.03 | (0.00, 0.06) | 0.076 |
| Externalizing (Level, Adjusted) |  | 4,236 | 0.00 | (-0.03, 0.03) | 0.939 | 3,948 | 0.01 | (-0.02, 0.04) | 0.519 |
| Internalizing (Level, Crude) |  | 4,238 | 0.02 | (-0.01, 0.05) | 0.130 | 3,950 | 0.03 | (0.00, 0.06) | 0.041 |
| Internalizing (Level, Adjusted) |  | 4,236 | 0.03 | (0.00, 0.06) | 0.084 | 3,948 | 0.04 | (0.01, 0.07) | 0.019 |
| Externalizing (Change, Crude) |  | 2,495 | 0.00 | (-0.04, 0.04) | 0.919 | 2,320 | -0.01 | (-0.05, 0.03) | 0.681 |
| Externalizing (Change, Adjusted) |  | 2,494 | 0.00 | (-0.04, 0.04) | 0.916 | 2,319 | -0.01 | (-0.05, 0.03) | 0.637 |
| Internalizing (Change, Crude) |  | 2,495 | -0.02 | (-0.05, 0.02) | 0.362 | 2,320 | -0.02 | (-0.06, 0.02) | 0.248 |
| Internalizing (Change, Adjusted) |  | 2,494 | -0.02 | (-0.06, 0.02) | 0.261 | 2,319 | -0.03 | (-0.06, 0.01) | 0.173 |

Abbreviations: CBCL = Child Behavior Checklist, N_obs_ = Number of observations

*Sensitivity Analyses – Adjusting for PDS Sum Score at Baseline as an Additional Covariate*

The Pubertal Development Scale (PDS) is a widely used, non-invasive questionnaire designed to assess pubertal status based on observable physical changes^38^. It includes items on body hair growth, skin changes, and growth spurts, as well as sex-specific items like breast development and menarche (for girls) or facial hair growth and voice deepening (for boys). Each item is rated on a 4-point Likert scale (1 = “has not begun” to 4 “already complete”), with the exception of menarche, which is binary (1 = no, 4 = yes). The final PDS score is calculated as the average across all items, providing a continuous measure of pubertal development.

In this study, we used the **parent-report version** of the PDS, which is particularly appropriate for younger participants^39^. For each participant, a **PDS Sum Score** was calculated as follows: First, responses were screened for validity. Participants with missing responses or non-informative answers (coded as “999”) on any of the five PDS items were excluded from the computation. Next, we computed the **mean across all five PDS items** separately for males and females (using sex-specific items). Finally, this mean score (ranging from 1 to 4) was multiplied by 5 to reconstruct a sum score equivalent to the total across all items, resulting in a PDS Sum Score ranging from 5 to 20. Higher scores indicate more advanced pubertal development.

Although we did not adjust for baseline PDS in the primary models to maintain consistency across hormones and maximize sample size, we performed a sensitivity analysis including it as an additional covariate to verify that associations between DHEA and mental health were not driven by pubertal stage.

**Supplementary Table S12. Sensitivity Analysis – Adjusting for PDS Sum Score at Baseline in Males: Association of DHEA Levels and CBCL at 3-Year Follow-Up.**

| **Outcome: CBCL Categories (Metric, Model)** |  | **Linear Mixed-Effects Models** | | | | **Negative Binominal Mixed Models** | | | |
| --- | --- | --- | --- | --- | --- | --- | --- | --- | --- |
|  |  | N_obs_ | b | 95%-CI | p | N_obs_ | b | 95%-CI | p |
| Externalizing (Level, Adjusted) |  | 4,676 | -0.07 | (-0.01, -0.04) | <0.001 | 4,526 | -0.07 | (-0.01, -0.04) | <0.001 |
| Internalizing (Level, Adjusted) |  | 4,676 | -0.03 | (-0.06, 0.00) | 0.022 | 4,526 | -0.04 | (-0.07, -0.01) | 0.017 |
| Externalizing (Change, Adjusted) |  | 2,733 | 0.01 | (-0.02, 0.05) | 0.500 | 2,660 | 0.01 | (-0.03, 0.05) | 0.547 |
| Internalizing (Change, Adjusted) |  | 2,733 | 0.00 | (-0.04, 0.04) | 0.962 | 2,660 | 0.00 | (-0.04, 0.04) | 0.987 |

Abbreviations: CBCL = Child Behavior Checklist, N_obs_ = Number of observations

**Supplementary Table S13. Sensitivity Analysis – Adjusting for PDS Sum Score at Baseline in Females: Association of DHEA Levels and CBCL at 3-Year Follow-Up.**

| **Outcome: CBCL Categories (Metric, Model)** |  | **Linear Mixed-Effects Models** | | | | **Negative Binominal Mixed Models** | | | |
| --- | --- | --- | --- | --- | --- | --- | --- | --- | --- |
|  |  | N_obs_ | b | 95%-CI | p | N_obs_ | b | 95%-CI | p |
| Externalizing (Level, Adjusted) |  | 4,236 | 0.00 | (-0.03, 0.03) | 0.939 | 4,107 | 0.00 | (-0.03, 0.03) | 0.959 |
| Internalizing (Level, Adjusted) |  | 4,236 | 0.03 | (0.00, 0.06) | 0.084 | 4,107 | 0.02 | (-0.01, 0.06) | 0.144 |
| Externalizing (Change, Adjusted) |  | 2,494 | 0.00 | (-0.04, 0.04) | 0.916 | 2,421 | 0.00 | (-0.03, 0.04) | 0.814 |
| Internalizing (Change, Adjusted) |  | 2,494 | -0.02 | (-0.06, 0.02) | 0.261 | 2,421 | -0.02 | (-0.05, 0.02) | 0.355 |

Abbreviations: CBCL = Child Behavior Checklist, N_obs_ = Number of observations

*Sensitivity Analyses - PDS Sum Score at 3-year Follow-Up instead of at baseline assessment*

The previous sensitivity analysis included the PDS Sum Score at baseline to account for pubertal status at study entry. However, pubertal development can change substantially between baseline and the 3-year follow-up, the time point at which the CBCL outcomes were assessed, potentially introducing additional variance. In our sample, the mean PDS Sum Score increased from 7.20 ± 1.85 in males and 8.88 ± 2.60 in females at baseline to 10.85 ± 3.18 in males and 14.63 ± 2.94 in females at the 3-year follow-up, reflecting substantial pubertal progression over this period.

To address this, we conducted a sensitivity analysis using the PDS Sum Score at the 3-year follow-up as a covariate. This allows for adjustment based on current pubertal status, which may more directly influence psychological symptoms. This sensitivity analysis therefore assesses the robustness of our findings when controlling for pubertal status at the outcome assessment, rather than only at baseline.

**Supplementary Table S14. Sensitivity Analysis – Adjusting for PDS Sum Score at 3-Year in Males: Association of DHEA Levels and CBCL at 3-Year Follow-Up.**

| **Outcome: CBCL Categories (Metric, Model)** |  | **Linear Mixed-Effects Models** | | | | | | | **Negative Binominal Mixed Models** | | | | | | |
| --- | --- | --- | --- | --- | --- | --- | --- | --- | --- | --- | --- | --- | --- | --- | --- |
|  |  | N_obs_ | | b | | 95%-CI | | p | N_obs_ | b | | | 95%-CI | | p |
| Externalizing (Level, Adjusted) |  | 4,676 | -0.07 | | (-0.01, -0.04) | | <0.001 | | 4,415 | | -0.07 | (-0.01, -0.04) | | <0.001 | |
| Internalizing (Level, Adjusted) |  | 4,676 | -0.03 | | (-0.06, 0.00) | | 0.022 | | 4,415 | | -0.04 | (-0.07, 0.00) | | 0.024 | |
| Externalizing (Change, Adjusted) |  | 2,733 | 0.01 | | (-0.02, 0.05) | | 0.500 | | 2,579 | | 0.01 | (-0.03, 0.05) | | 0.621 | |
| Internalizing (Change, Adjusted) |  | 2,733 | 0.00 | | (-0.04, 0.04) | | 0.962 | | 2,579 | | -0.01 | (-0.05, 0.03) | | 0.699 | |

Abbreviations: CBCL = Child Behavior Checklist, N_obs_ = Number of observations

**Supplementary Table S15. Sensitivity Analysis – Adjusting for PDS Sum Score at 3-Year in Females: Association of DHEA Levels and CBCL at 3-Year Follow-Up.**

| **Outcome: CBCL Categories (Metric, Model)** |  | **Linear Mixed-Effects Models** | | | | | | **Negative Binominal Mixed Models** | | | | |
| --- | --- | --- | --- | --- | --- | --- | --- | --- | --- | --- | --- | --- |
|  |  | N_obs_ | | b | 95%-CI | | p | N_obs_ | b | | 95%-CI | p |
| Externalizing (Level, Adjusted) |  | 4,236 | 0.00 | | | (-0.03, 0.03) | 0.939 | 3,883 | -0.01 | (-0.05, 0.02) | | 0.458 |
| Internalizing (Level, Adjusted) |  | 4,236 | 0.03 | | | (0.00, 0.06) | 0.084 | 3,883 | 0.01 | (-0.02, 0.05) | | 0.461 |
| Externalizing (Change, Adjusted) |  | 2,494 | 0.00 | | | (-0.04, 0.04) | 0.916 | 2,292 | -0.01 | (-0.05, 0.03) | | 0.660 |
| Internalizing (Change, Adjusted) |  | 2,494 | -0.02 | | | (-0.06, 0.02) | 0.261 | 2,292 | -0.03 | (-0.06, 0.01) | | 0.180 |

Abbreviations: CBCL = Child Behavior Checklist, N_obs_ = Number of observations

*Sensitivity Analyses – Excluding Participants with Hormone Levels below Lower Limit of Detection Instead of Imputation.*

In the main adjusted models, hormone values below the lower limit of detection (LLD) were imputed to preserve statistical power and reduce bias due to non-random missingness. However, imputation introduces assumptions about the distribution of undetectable values, which cannot directly be tested and may influence the results.

To test the robustness of our findings, we conducted a sensitivity analysis excluding all participants with DHEA levels below the LLD, thus relying exclusively on observed hormone values. This approach eliminates potential biases introduced by imputation and provides complementary evidence for the stability of associations between DHEA and CBCL outcomes.

**Supplementary Table S16. Sensitivity Analysis – Excluding Males with Hormone Levels below LLD: Association of Mean DHEA Levels at 3-Year Follow-Up.**

| **Outcome: CBCL Categories (Metric, Model)** |  | **Primary Analysis: Values below LLD imputed** | | | | **Sensitivity Analysis: Values below LLD excluded** | | | |
| --- | --- | --- | --- | --- | --- | --- | --- | --- | --- |
|  |  | N_obs_ | b | 95%-CI | p | N_obs_ | b | 95%-CI | p |
| Externalizing (Level, Crude) |  | 4,676 | -0.06 | (-0.09, -0.03) | <0.001 | 4,670 | -0.06 | (-0.09, -0.03) | <0.001 |
| Externalizing (Level, Adjusted) |  | 4,676 | -0.07 | (-0.10, -0.04) | <0.001 | 4,670 | -0.07 | (-0.10, -0.04) | <0.001 |
| Internalizing (Level, Crude) |  | 4,676 | -0.03 | (-0.06, 0.00) | 0.064 | 4,670 | -0.03 | (-0.06, 0.00) | 0.078 |
| Internalizing (Level, Adjusted) |  | 4,676 | -0.03 | (-0.06, 0.00) | 0.022 | 4,670 | -0.03 | (-0.06, 0.00) | 0.027 |
| Externalizing (Change, Crude) |  | 2,733 | 0.01 | (-0.02, 0.05) | 0.453 | 2,730 | 0.01 | (-0.02, 0.05) | 0.439 |
| Externalizing (Change, Adjusted) |  | 2,733 | 0.01 | (-0.02, 0.05) | 0.500 | 2,730 | 0.01 | (-0.02, 0.05) | 0.487 |
| Internalizing (Change, Crude) |  | 2,733 | 0.00 | (-0.04, 0.04) | 0.943 | 2,730 | 0.00 | (-0.04, 0.04) | 0.950 |
| Internalizing (Change, Adjusted) |  | 2,733 | 0.00 | (-0.04, 0.04) | 0.962 | 2,730 | 0.00 | (-0.04, 0.04) | 0.954 |

Abbreviations: LLD = Lower Limit of Detection, CBCL = Child Behavior Checklist, N_obs_ = Number of observations

**Supplementary Table S17. Sensitivity Analysis – Excluding Females with Hormone Levels below LLD: Association of Mean DHEA Levels at 3-Year Follow-Up.**

| **Outcome: CBCL Categories (Metric, Model)** |  | **Primary Analysis: Values below LLD imputed** | | | | **Sensitivity Analysis: Values below LLD excluded** | | | |
| --- | --- | --- | --- | --- | --- | --- | --- | --- | --- |
|  |  | N_obs_ | b | 95%-CI | p | N_obs_ | b | 95%-CI | p |
| Externalizing (Level, Crude) |  | 4,238 | 0.02 | (-0.01, 0.05) | 0.290 | 4,236 | 0.02 | (-0.01, 0.05) | 0.285 |
| Externalizing (Level, Adjusted) |  | 4,236 | 0.00 | (-0.03, 0.03) | 0.939 | 4,234 | 0.00 | (-0.03, 0.03) | 0.942 |
| Internalizing (Level, Crude) |  | 4,238 | 0.02 | (-0.01, 0.05) | 0.130 | 4,236 | 0.03 | (0.00, 0.06) | 0.100 |
| Internalizing (Level, Adjusted) |  | 4,236 | 0.03 | (0.00, 0.06) | 0.084 | 4,234 | 0.03 | (0.00, 0.06) | 0.064 |
| Externalizing (Change, Crude) |  | 2,495 | 0.00 | (-0.04, 0.04) | 0.919 | 2,493 | 0.00 | (-0.04, 0.04) | 0.922 |
| Externalizing (Change, Adjusted) |  | 2,494 | 0.00 | (-0.04, 0.04) | 0.916 | 2,492 | 0.00 | (-0.04, 0.04) | 0.922 |
| Internalizing (Change, Crude) |  | 2,495 | -0.02 | (-0.05, 0.02) | 0.362 | 2,493 | -0.02 | (-0.05, 0.02) | 0.382 |
| Internalizing (Change, Adjusted) |  | 2,494 | -0.02 | (-0.06, 0.02) | 0.261 | 2,492 | -0.02 | (-0.06, 0.02) | 0.278 |

Abbreviations: LLD = Lower Limit of Detection, CBCL = Child Behavior Checklist, N_obs_ = Number of observations

*Sensitivity Analyses – Adjusting for Baseline CBCL Scores.*

It can be assumed that earlier symptomatology and CBCL scores at baseline are predictive of higher scores at later time points. Therefore, we ran an expanded model that included baseline CBCL scores as an additional covariate. The results yielded similar results, however, with slightly attenuated effect sizes across all follow-up assessments.

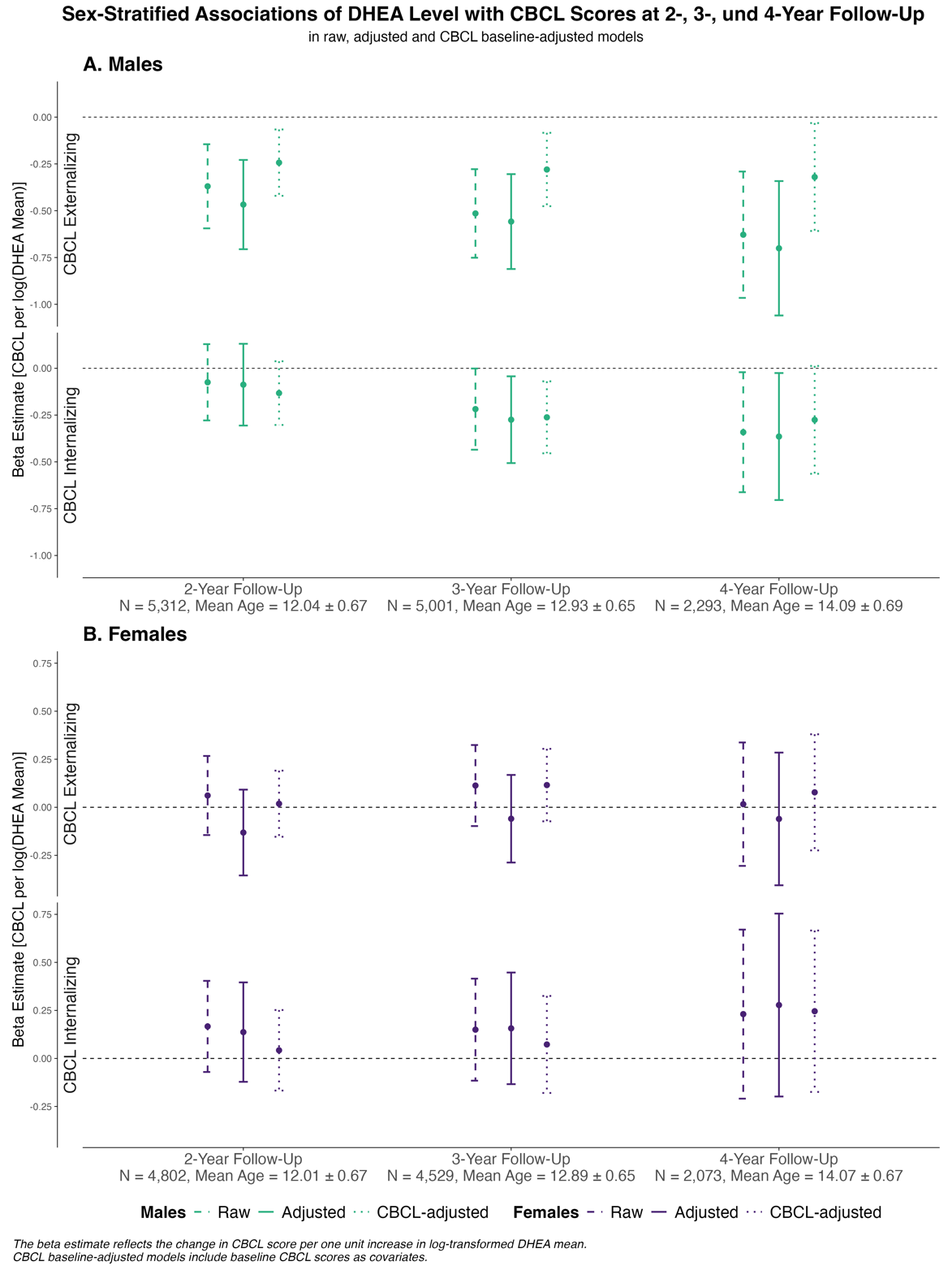

**Supplementary Figure S15. Sensitivity Analysis – Sex-Stratified Associations Between DHEA Levels and CBCL Scores Across Follow-Ups in Raw, Adjusted, and Baseline-CBCL-Adjusted Models.** Panel A: Males, Panel B: Females. Beta estimates (points) and 95% confidence intervals (vertical lines) are shown for three model specifications: raw (dashed line), adjusted for covariates (solid line), and additionally adjusted for baseline CBCL score (dotted line). Raw models include only baseline age and a random intercept for family nested within study site. Adjusted models additionally include the minimally sufficient adjustment set: baseline values of age, race/ethnicity, BMI standard deviation score, physical activity, and the random intercept for family nested within study site. Baseline-CBCL-adjusted models further control for baseline CBCL scores. Beta estimates reflect the change in CBCL score per one unit increase in log-transformed mean DHEA level. Sample sizes and mean ages for each follow-up are reported below.

**Supplementary Results**

*Correlations between Hormone Levels across Timepoints, Unadjusted and Adjusted for Current Pubertal Stage (PDS).*

Correlations between hormone levels across timepoints were calculated using Pearson’s correlation. To account for pubertal status, linear regression models including the PDS score as a covariate were fitted. Adjusted regression coefficients and p-values are reported, providing estimates of hormone associations independent of PDS.

**Supplementary Table S18. Correlations between Hormones across Timepoints.**

|  |  | **Males** | | | **Females** | | | | | |
| --- | --- | --- | --- | --- | --- | --- | --- | --- | --- | --- |
|  |  | **N** | **r** | **p** | **N** | | **r** | | | **p** |
|  | **DHEA – Testosterone** | | | | | | | | |  |
|  | Baseline | 4,554 | 0.759 | <0.001 | 4,161 | | 0.809 | | | <0.001 |
|  | 1-year | 4,753 | 0.756 | <0.001 | 4,372 | | 0.803 | | | <0.001 |
|  | 2-year | 5,038 | 0.752 | <0.001 | 4,596 | | 0.799 | | | <0.001 |
|  | 3-year | 4,742 | 0.751 | <0.001 | 4,336 | | 0.799 | | | <0.001 |
|  | 4-year | 2,168 | 0.748 | <0.001 | 1,971 | | 0.794 | | | <0.001 |
|  | **DHEA – Testosterone PDS adjusted** | | | | | |  | | |  |
|  | Baseline | 4,222 | 0.759 | <0.001 | 3,870 | | 0.807 | | | <0.001 |
|  | 1-year | 4,428 | 0.758 | <0.001 | 4,067 | | 0.803 | | | <0.001 |
|  | 2-year | 4,679 | 0.753 | <0.001 | 4,212 | | 0.799 | | | <0.001 |
|  | 3-year | 4,396 | 0.751 | <0.001 | 3,876 | | 0.796 | | | <0.001 |
|  | 4-year | 2,040 | 0.743 | <0.001 | 1,780 | | 0.793 | | | <0.001 |
|  | **DHEA – Estradiol** | | | | | | |  | | |
|  | Baseline |  |  |  | 4,135 | | 0.552 | | | <0.001 |
|  | 1-year |  |  |  | 4,330 | | 0.560 | | | <0.001 |
|  | 2-year |  |  |  | 4,547 | | 0.549 | | | <0.001 |
|  | 3-year |  |  |  | 4,286 | | 0.547 | | | <0.001 |
|  | 4-year |  |  |  | 1,949 | | 0.536 | | | <0.001 |
| **DHEA – Estradiol PDS adjusted** | | | | | |  | | |  |  |
| Baseline | |  |  |  | 3,845 | | 0.554 | | | <0.001 |
| 1-year | |  |  |  | 4,028 | | 0.558 | | | <0.001 |
| 2-year | |  |  |  | 4,171 | | 0.547 | | | <0.001 |
| 3-year | |  |  |  | 3,834 | | 0.544 | | | <0.001 |
| 4-year | |  |  |  | 1,761 | | 0.538 | | | <0.001 |

Abbreviations: N = Number; p = p-value; PDS = Pubertal Development Scale; r = Pearson’s R Correlation Coefficient.

*Testosterone as an Additional Exposure to Assess Incremental Variance Explained*

To assess whether testosterone contributes additional explanatory power beyond DHEA, we conducted analyses including testosterone as an additional predictor in the 3-year mixed-effects models. In these models, marginal R² represents the proportion of variance explained by the fixed effects alone, while conditional R² reflects the variance explained by both fixed and random effects. This distinction allows to separate the contribution of hormone predictors from the overall variance captured by the model structure.

For CBCL externalizing scores, the 3-year models including DHEA alone showed a marginal R² of 0.010 and conditional R² of 0.389 in males (p = <0.001), and a marginal R² of 0.010 and conditional R² of 0.513 in females (p = 0.939). Adding testosterone did not meaningfully change model fit (males: marginal R² = 0.010, conditional R² = 0.385, p = <0.001; females: marginal R² = 0.010, conditional R² = 0.513, p = 0.671). For CBCL internalizing scores, the DHEA-only models showed a marginal R² of 0.016 and conditional R² of 0.569 in males (p = 0.022), and a marginal R² of 0.018 and conditional R² of 0.687 in females (p = 0.084). Including testosterone resulted in minimal changes (males: marginal R² = 0.016, conditional R² = 0.565, p = 0.087; females: marginal R² = 0.018, conditional R² = 0.685, p = 0.066).

These results indicate that testosterone does not explain additional variance in CBCL externalizing or internalizing scores beyond DHEA. This finding supports the interpretation that the observed hormone-psychopathology associations primarily reflect adrenal rather than gonadal contributions.

*Analysis of DSM5-Oriented CBCL Subdomains Across Follow-Up Years*

While the primary focus was on the CBCL categories of Externalizing and Internalizing symptoms based on the syndrome-oriented scales, we further examined associations between DHEA levels and changes with the six DSM-5-oriented CBCL subscales, which align more closely with clinical diagnostic categories. These subscales included Attention-Deficit/Hyperactivity Disorder (ADHD), Oppositional Defiant Disorder (ODD), and Conduct Disorder (CD) for Externalizing symptoms, as well as Depression, Anxiety, and Somatic Problems for Internalizing symptoms. The results indicate that the main findings are reflected across all CBCL subscores. One exception was ADHD in girls at the 2-year follow-up, which showed a negative estimate (β = –0.17, 95% CI [–0.28, –0.06]). However, this association was not observed at later follow-ups.

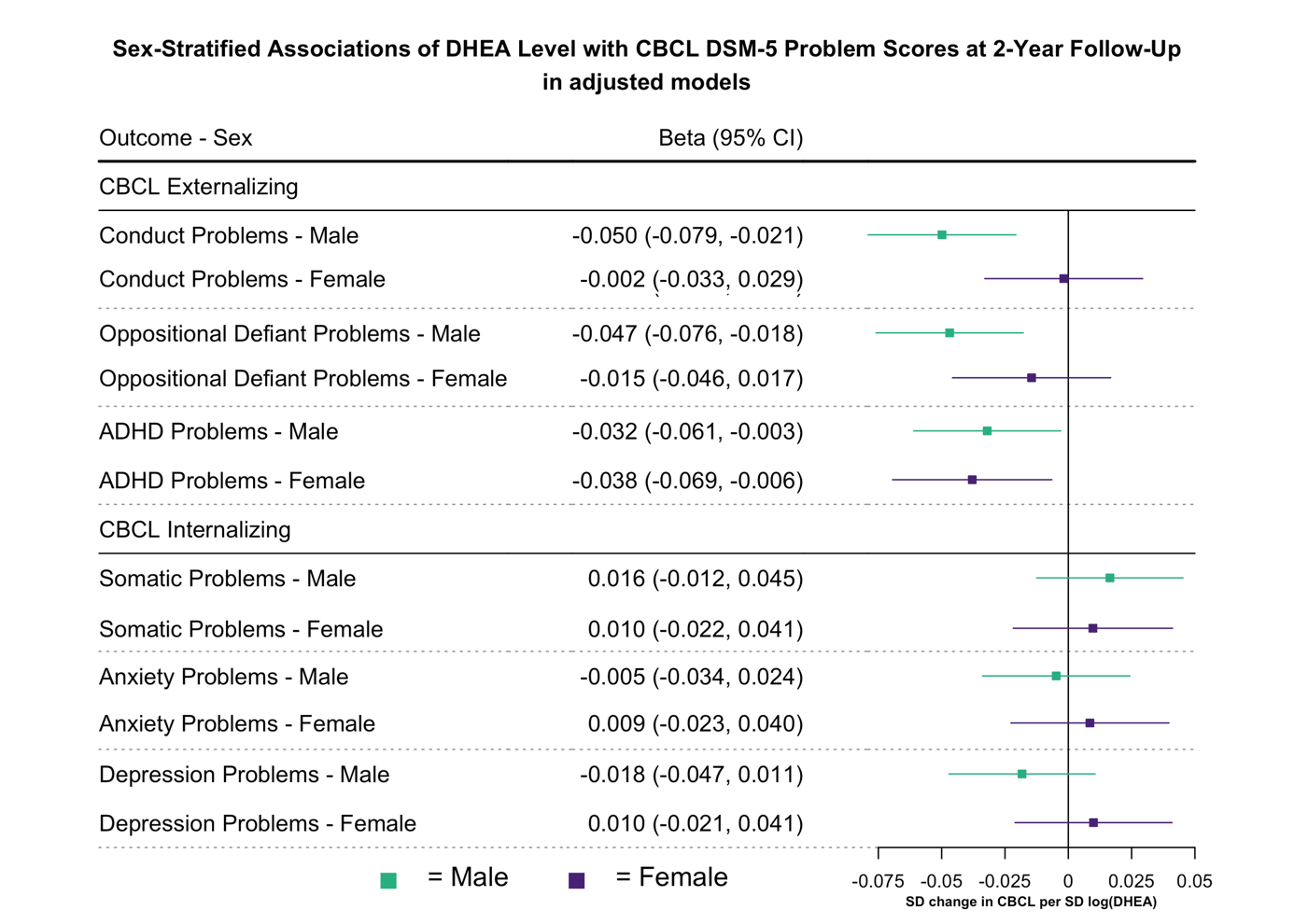

**Supplementary Figure S16. Associations of DHEA with DSM-5 Problem Scores at 2-Year Follow-Up.**

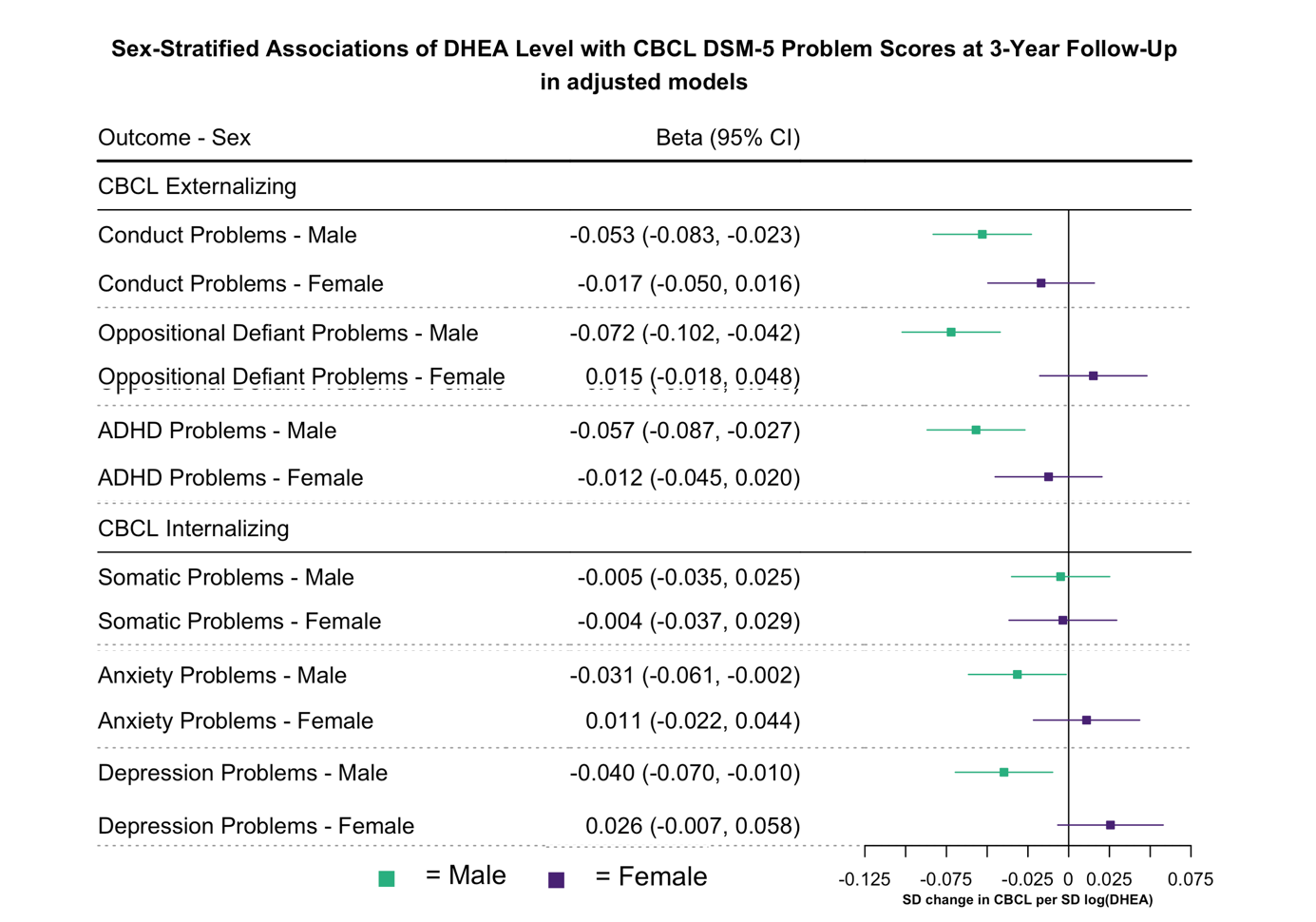

**Supplementary Figure S17. Associations of DHEA with DSM-5 Problem Scores at 3-Year Follow-Up.**

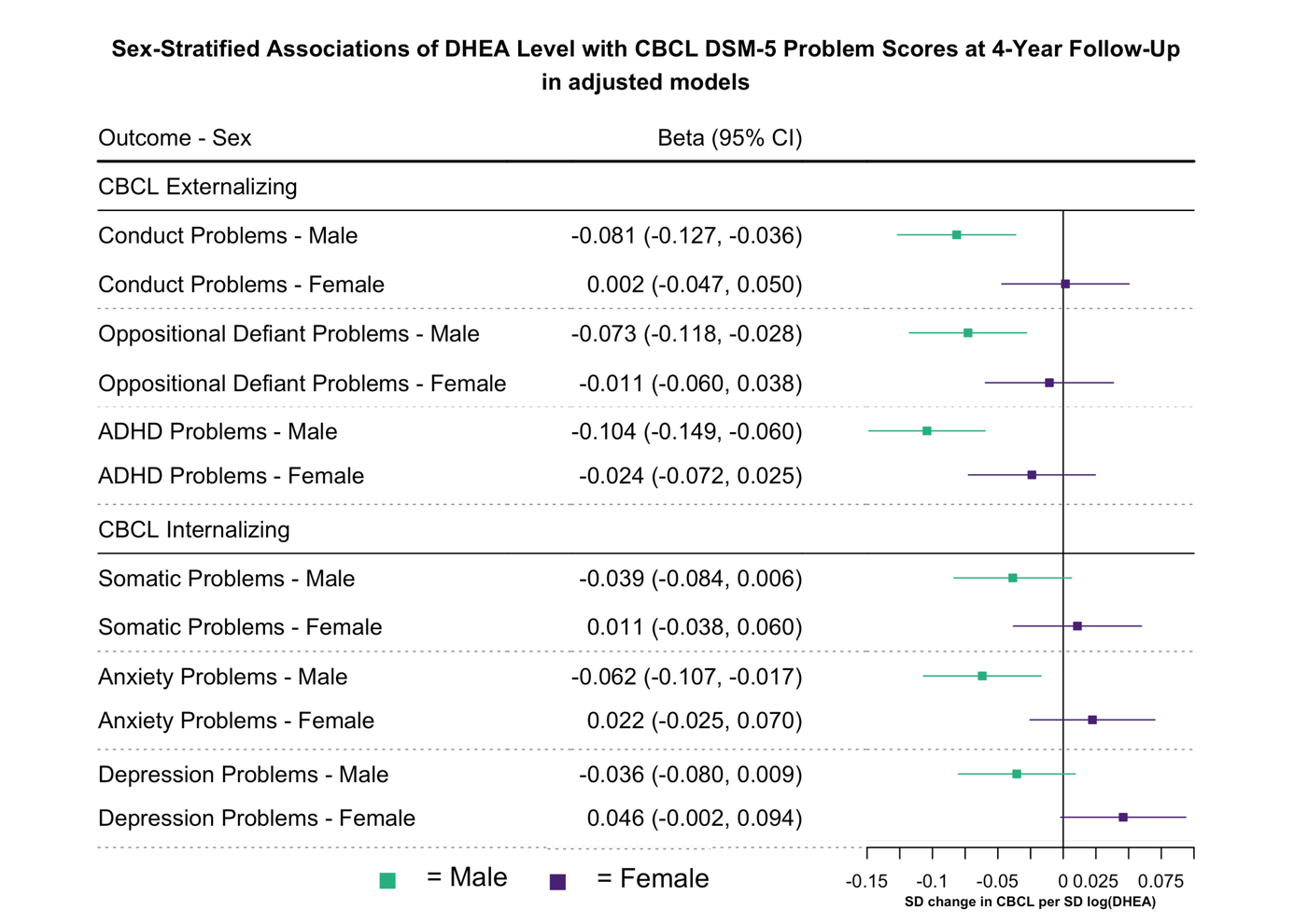

**Supplementary Figure S18. Associations of DHEA with DSM-5 Problem Scores at 4-Year Follow-Up.**

*Risk Ratio Analysis*

To improve interpretability of the main analysis results, we computed risk ratios (RRs) for the association between DHEA levels and CBCL outcomes at 2-, 3-, and 4-year follow-up assessments. Therefore, we estimated adjusted risk ratios based on binomial regression models with a log link, directly quantifying the **relative change in risk** for meeting CBCL-defined thresholds per one-unit increase in log-transformed DHEA levels. These models were adjusted for age, race/ethnicity, BMI-SDS and physical activity, all assessed at baseline and were conducted separately for males and females. Due to convergence issues, random intercepts for family and site were not included in the final models. As a result, models do not account for potential clustering at the family or site level.

CBCL outcomes were categorized based on established T-score thresholds:

- T-score ≤ 59: Non-pathological (normal range)
- T-score 60–63: Borderline range
- T-score ≥ 64: Clinically significant (clinical range)

Separate models were computed for Externalizing and Internalizing problems at each follow-up time point (2, 3, and 4 years). For each domain, two binary outcome definitions were applied:

1. Borderline + Clinical**:** T-score ≥ 60 vs. T-score ≤ 59
2. Clinical: T-score ≥ 64 vs. T-score ≤ 59

This approach yielded four outcome models per follow-up year, allowing for consistent comparison of adjusted risk estimates across symptom domains and time points. Results are presented in Figure 2.

*Sex-Interaction Effects*

To investigate whether the relationship between hormonal levels and mental health outcomes varied by sex, we fitted linear mixed-effects models including a sex-by-hormone interaction term. We compared four nested models: (1) a baseline model with sex and covariates only, (2) a main-effects model adding hormone exposure (log-transformed mean or change in DHEA), and (3) a full model incorporating the sex × hormone interaction and (4) a model incorporating the sex × hormone interaction with CBCL externalizing/internalizing scores assessed at baseline as an additional covariate as prior symptomatology is known to predict later CBCL scores.

The sex-interaction effects of DHEA level on CBCL outcomes are illustrated in Figure 1. The table below further presents the effects of DHEA change across follow-ups, which did not show consistent or meaningful differences. We report beta estimates and 95% confidence intervals for our models. Adjusting for baseline CBCL scores did not substantially alter the estimates.

**Supplementary Table S19. DHEA × Sex-Interaction Effects on CBCL Outcomes Across Follow-Up Years.**

| **DHEA**  **Exposure** | **CBCL Outcome** | **Sex Effect** | | **Sex Interaction** | | **CBCL baseline** ^c^ | |
| --- | --- | --- | --- | --- | --- | --- | --- |
|  |  | **b** | **95%-CI** | **b** | **95%-CI** | **b** | **95%-CI** |
| **2-Year Follow-Up** | | | | | | | |
| Level ^a^ | Externalizing | -0.15 | (-0.19, -0.11) | 0.05 | (0.01, 0.09) | 0.05 | (0.02, 0.08) |
|  | Internalizing | 0.10 | (0.05, 0.14) | 0.04 | (0.01, 0.08) | 0.04 | (0.01, 0.07) |
| Change ^b^ | Externalizing | -0.17 | (-0.22, -0.11) | -0.03 | (-0.08, 0.02) | -0.02 | (-0.06, 0.02) |
|  | Internalizing | 0.09 | (0.04, 0.14) | -0.03 | (-0.08, 0.03) | 0.00 | (-0.04, 0.04) |
| **3-Year Follow-Up** | |  |  |  |  |  |  |
| Level ^a^ | Externalizing | -0.15 | (-0.19, -0.11 ) | 0.07 | (0.03, 0.11) | 0.06 | (0.03, 0.09) |
|  | Internalizing | 0.17 | (0.13, 0.21) | 0.05 | (0.01, 0.09) | 0.05 | (0.01, 0.08) |
| Change ^b^ | Externalizing | -0.16 | (-0.22, -0.11) | -0.01 | (-0.07, 0.04) | 0.00 | (-0.04, 0.04) |
|  | Internalizing | 0.17 | (0.12, 0.23) | -0.02 | (-0.07, 0.03) | 0.01 | (-0.04, 0.05) |
| **4-Year Follow-Up** | |  |  |  |  |  |  |
| Level ^a^ | Externalizing | -0.09 | (-0.16, -0.03) | 0.08 | (0.01, 0.14) | 0.04 | (0.01, 0.10) |
|  | Internalizing | 0.27 | (0.21, 0.33) | 0.08 | (0.02, 0.14) | 0.05 | (0.00, 0.11) |
| Change ^b^ | Externalizing | -0.12 | (-0.20, -0.05) | -0.01 | (-0.08, 0.06) | 0.00 | (-0.06, 0.06) |
|  | Internalizing | 0.27 | (0.20, 0.34) | -0.04 | (-0.10, 0.03) | -0.01 | (-0.07, 0.05) |

Abbreviations: DHEA = Dehydroepiandrosterone; CBCL = Child Behavior Checklist; b = Beta Estimate; 95%-CI = 95% Confidence Interval

a DHEA Level is defined as the natural logarithm of the arithmetic mean of Baseline and 1-Year Follow-Up DHEA values per participant.

b DHEA Change is defined as the natural logarithm of the ratio between 2-Year Follow-Up and Baseline DHEA values per participant.

c Sex-interaction model with CBCL baseline score as additional covariate.

Models were estimated using linear mixed-effects modeling (lmer package in R).
